# Cross-ancestry genetic architecture and prediction for cholesterol traits

**DOI:** 10.1101/2023.01.31.23285307

**Authors:** Md. Moksedul Momin, Xuan Zhou, Elina Hyppönen, Beben Benyamin, S. Hong Lee

**Affiliations:** Australian Centre for Precision Health, University of South Australia, Adelaide, SA, 5000, Australia; UniSA Allied Health and Human Performance, University of South Australia, Adelaide, SA, 5000, Australia; Department of Genetics and Animal Breeding, Faculty of Veterinary Medicine, Chattogram Veterinary and Animal Sciences University (CVASU), Khulshi, Chattogram, 4225, Bangladesh; South Australian Health and Medical Research Institute (SAHMRI), University of South Australia, Adelaide, SA, 5000, Australia; UniSA Clinical and Health Sciences, University of South Australia, Adelaide, SA, Australia

## Abstract

While cholesterol is essential for human life, a high level of cholesterol is closely linked with the risk of cardiovascular diseases. Genome-wide association studies (GWASs) have been successful to identify genetic variants associated with cholesterol, which have been conducted mostly in white European populations. Consequently, it remains mostly unknown how genetic effects on cholesterol vary across ancestries. Here, we estimate cross-ancestry genetic correlation to address questions on how genetic effects are shared across ancestries for cholesterol. We find significant genetic heterogeneity between ancestries for total- and LDL-cholesterol. Furthermore, we show that single nucleotide polymorphisms (SNPs), which have concordant effects across ancestries for cholesterol, are more frequently found in the regulatory region, compared to the other genomic regions. Indeed, the positive genetic covariance between ancestries is mostly driven by the effects of the concordant SNPs, whereas the genetic heterogeneity is attributed to the discordant SNPs. We also show that the predictive ability of the concordant SNPs is significantly higher than the discordant SNPs in the cross-ancestry polygenic prediction. The list of concordant SNPs for cholesterol is available in GWAS Catalog (https://www.ebi.ac.uk/gwas/; details are in web resources section). These findings have relevance for the understanding of shared genetic architecture across ancestries, contributing to the development of clinical strategies for polygenic prediction of cholesterol in cross-ancestral settings

## Introduction

Cholesterol is a type of lipid that is essential for human life, forming an essential structural component of the cell membrane^1-3^. While cholesterol is necessary for human body to function, too much cholesterol can harm the body. High cholesterol is linked with a high risk of cardiovascular diseases (CVDs), such as coronary heart disease, stroke, and peripheral vascular disease, which are the leading cause of death worldwide^4^, accounting for 32% of all deaths in 2019^5^. Specifically, elevated low-density lipoprotein (LDL) and decreased high-density lipoprotein (HDL) cholesterols are associated with increased CVD risk^6-9^. These cholesterol traits are heritable and known to be polygenic^6, 10, 11^. Reported heritability estimates for total-, LDL- and HDL-cholesterols are typically in the range of 20 to 60%^12^.

Over the last two decades, genome-wide association studies (GWASs) have successfully identified several genome-wide significant single nucleotide polymorphisms (SNPs) associated with cholesterol traits^4, 13-15^. While these findings have provided important insights into the genetics of cholesterol, most GWAS for cholesterol to date have been conducted in populations of white European ancestry^16-18^. Although the number of GWASs representing non-European populations are gradually increasing, they still remain greatly underrepresented in the efforts of gene discovery^16, 19^. Consequently, how genetic effects on cholesterol vary across ancestries remain mostly unknown^20, 21^. It is also not clear to what extent the associated genetic variants discovered in European populations are relevant for other ancestries (e.g., South Asian and African ancestries), and if the polygenic risk prediction of cholesterol can be applied across ancestries^22-25^.

The genetic effects on most complex traits are likely to vary at least to some extent across different ancestry groups^26, 27^. Cross-ancestry genetic correlation analyses can dissect the shared genetic architecture between diverse ancestries, also allowing to leverage power from diverse sources of information^28^. While common causal variants for cholesterol are likely to be shared across ancestries, their per-allele effect sizes may depend on allele frequencies that can differ across ancestries due to different evolutionary force such as selection and genetic drift^29^. Moreover, each ancestry has a unique genetic background that may affect the magnitude and direction of per-allele effect sizes for complex traits such as cholesterol^30^. It has been reported that the relationship between allele frequency and per-allele effect size varies across different ancestries, which should be properly accounted for. otherwise, the estimation of cross-ancestry genetic correlation can be biased^31, 32^.

Cross-ancestry genetic prediction can reduce the potential health disparity for non-European populations that are still underrepresented in public genomic databases including GWAS and polygenic risk scores (PRS)^33^. It is crucial to understand the source of genetic heterogeneity across ancestries in the genetic prediction. In general, it is not likely that SNP effects estimated from a single ancestry group are always applicable to other ancestries, which has a practical relevance. For example, several studies have reported that the predictive ability of complex traits including cholesterol was poor for Africans, East-Asians, South-Asians and Latinos, when using SNP effects estimated in Europeans^19, 34, 35^. To obtain more reliable cross-ancestry genetic prediction, it may be important to restrict to functionally homogenous genes or common causal variants across ancestries^28, 36^. We hypothesize that SNPs in strong linkage disequilibrium (LD) with the functionally homogenous genes have concordant effects, i.e., the same direction of SNP effects, across ancestries.

In this study, we estimate cross-ancestry genetic correlation to address the question about how genetic effects are shared across ancestries for cholesterol traits, accounting for the relationship between allele frequency and per-allele effect size^31^. In the estimation of cross-ancestry genetic correlation, we also investigate the role of concordant SNPs that are derived from comparing SNP effects between two independent GWAS datasets of UK Biobank and Biobank Japan (BBJ). We evaluate the transferability of genetic prediction across different ancestry groups and suggest a list of SNPs that are suitable for the use in polygenic risk prediction in cross-ancestry analyses.

## Results

### Overview of methods

The total numbers of individuals and SNPs for each ancestry after stringent quality control (QC) (Methods) are shown in **Supplementary Table 1**. From the quality-controlled data of 288,837 white British people, we randomly selected 30,000 individuals to be used in the analyses of cross-ancestry genetic correlations. The remaining 258,792 individuals were used as the discovery dataset in the cross-ancestry genetic risk prediction and in the classification of concordant SNPs (referred to as UKBB discovery). In the cross-ancestry genetic analysis of total-, HDL- and LDL-cholesterol, four ancestry groups were included, i.e. the 30,000 white British ancestry group, 26,457 other European, 6,199 south Asian and 6,179 African ancestry groups (**Supplementary Table 1**). We accounted for the relationship between per-allele effect size and allele frequency^31, 37^ by using trait-specific and ancestry-specific *α* that was explicitly estimated for each trait and each ancestry, using Akaike information criterion (AIC)^31, 32^. We used the common SNPs for each pair of ancestries to estimate the cross-ancestry genetic correlation, using the bivariate GREML approach^38^, accounting for the relationship between allele frequency and per-allele effect size^31^. We further investigated if the set of concordant SNPs, which were derived by comparing UKBB and Biobank Japan discovery GWAS summary statistics for cholesterol, is enriched in the regulatory region, compared to the other genomic regions. The list of concordant SNPs for total-, HDL- and LDL-cholesterol are now available in GWAS catalogue. Cross-ancestry genetic covariance was partitioned, based on the sets of concordant and discordant SNPs, to see how the genetic heterogeneity is attributed to those SNP sets (see **Supplementary Table 4)**. Finally, cross-ancestry polygenic prediction was performed based on the sets of concordant and discordant SNPs.

### Determining trait-specific and ancestry-specific scale factor (*α*) for each ancestry

The scale factor (*α*) can account for the relationship between allele frequency and per-allele effect size, that is, per-allele effect sizes vary, proportional to [*p* (1 − *p*)] ^*α*^, where *p* is the allele frequency^32, 39, 40^. It is also reported that the scale factor is not uniformly distributed across ancestries, and there may be an optimal *α* value for each specific ancestry group^31^. Following the previous approach^31, 32^, we investigated various *α* values ranging between -1 and 0.5 to determine the ancestry specific *α* value of each ancestry group for total-, LDL- and HDL-cholesterol. To determine optimal *α*, we compared the Akaike Information Criteria (AIC) values across different heritability models with various *α* values for each trait and each ancestry **(Figure 1)**. Detailed values of log-likelihood and AIC are provided in **Supplementary Table 6-9**. As expected, optimal *α* values are not uniformly distributed across traits and across ancestries (Figure 1). These identified *α* values are subsequently used in the estimation of cross-ancestry genetic correlations to dissect the shared genetic architecture and investigate genetic heterogeneity across ancestries for the cholesterol related traits.

**Figure 1:**
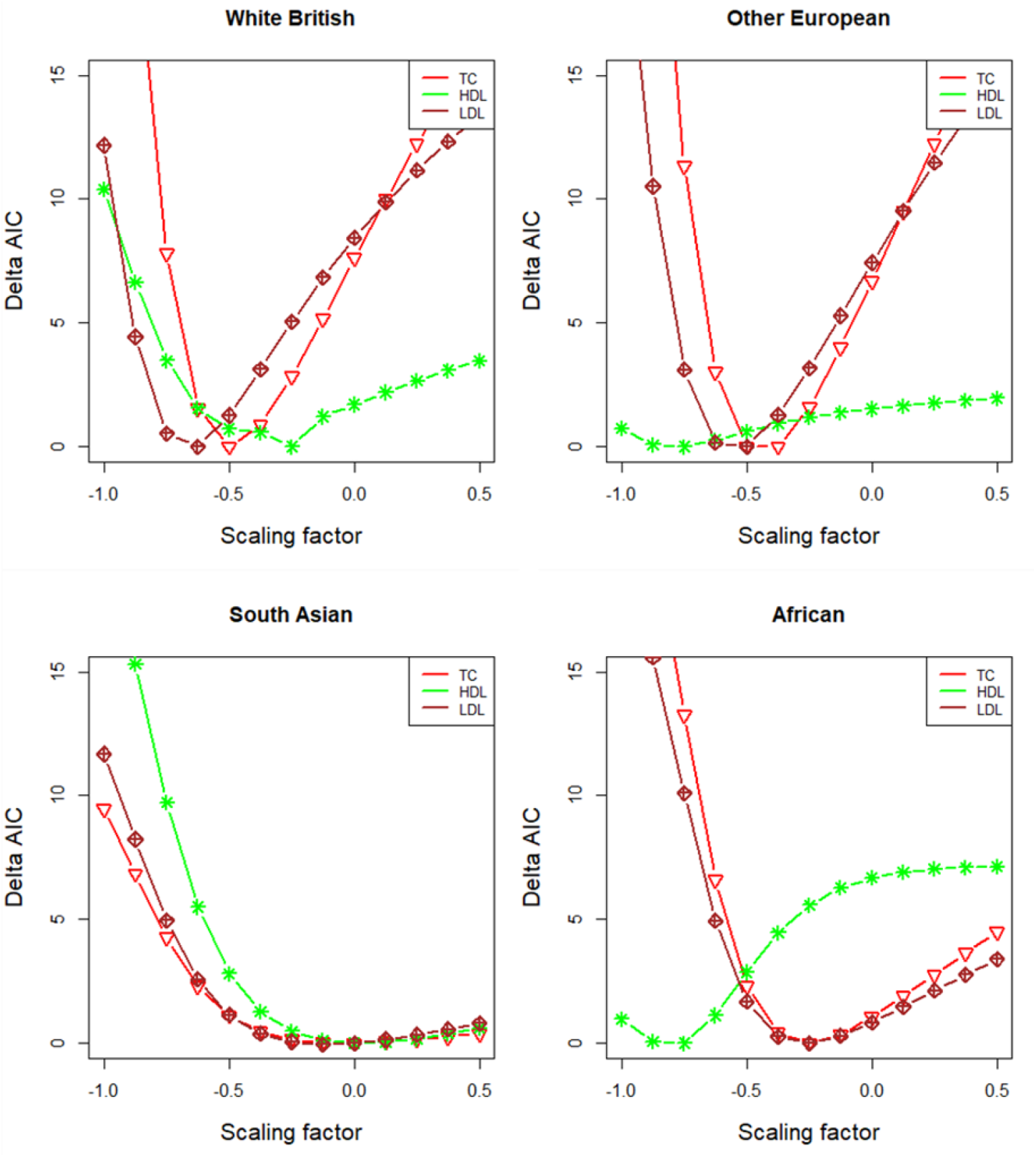
Determining the optimal ancestry-specific scaling factors (*α*) for each trait. The *α* value reflects the relationship between allele frequency and per-allele effect size and can vary across ancestries and traits. ΔAIC values are plotted against scaling factors, *α*, for each ancestry group. The lowest AIC (i.e., ΔAIC=0) indicates the best model. The sample sizes are 30,000, 26,457, 6,199, and 6,179 for white British, other European, South Asian, and African ancestry groups, respectively. TC: total-cholesterol, HDL: high-density lipoprotein cholesterol, LDL: low-density lipoprotein cholesterol.

### Heritability (*h*^*2*^) estimates across ancestries

The estimated SNP-based heritabilities of total-, LDL- and HDL-cholesterol are presented in Figure 1. The estimates are significantly different particularly between European and African ancestries. For total-cholesterol, there is a significant difference in SNP-based heritability estimates between African vs. European (*p-*value=4.26e-03), and African vs white British (*p-* value=1.14e-03). Similarly, the estimate of LDL-cholesterol is significantly lower in white British (*p-*value= 1.11e-03) and other European (*p-*value= 5.19e-03) than African ancestry, which agrees with the previous findings based on twin studies^41^. We also observed significant heterogeneity of SNP-based heritability for the HDL-cholesterol between South Asian and other Europeans, between South Asian and white British.

### Estimated cross-ancestry genetic correlations

The estimated cross-ancestry genetic correlations (*r*_*g*_) for cholesterol traits are presented in **Figure 3**. For total-cholesterol, we observed a genetic heterogeneity between South Asian vs. white British (*r*_*g*_= 0.399; SE= 0.143; *p*-value= 2.65e-05), South Asian vs. other European (*r*_*g*_= 0.353; SE=0.133; *p*-value= 1.14e-06) and South Asian vs. African ancestry (*r*_*g*_ = 0.188; SE=0.197; *p*-value= 3.76e-05). There is also a genetic heterogeneity between African vs. white British (*r*_*g*_= 0.473; SE=0.127; *p*-value= 3.33e-05) and African vs. other European ancestry (*r*_*g*_= 0.315; SE=0.122; *p*-value=1.96e-08). In contrast, white British and other European are genetically homogenous (*r*_*g*_ =0.954; SE=0.087; *p*-value= 5.96e-01) (**Figure 3 and Supplementary Table 10**). For LDL-cholesterol, results are similar to total-cholesterol. There is a significant genetic heterogeneity between South Asian vs. white British (*r*_*g*_ = 0.296; SE=0.155; *p*-value=5.57e-06), South Asian vs. other European (*r*_*g*_ = 0.177; SE=0.138; *p*-value=2.46e-09), South Asian vs. African (*r*_*g*_ = 0.110; SE=0.190; *p*-value=2.81e-06), and African vs. other European ancestry (*r*_*g*_= 0.409; SE=0.147; *p*-value=2.81e-06) (**Figure 3 and Supplementary Table 11**). As expected, the cross-ancestry genetic correlation between other European and white British was close to 1 (*r*_*g*_= 1.084; SE=0.128; *p*-value=5.12e-01). We did not observe genetic heterogeneity among the pairs of ancestry groups for HDL-cholesterol (**Figure 3 and Supplementary Table 12)**.

**Figure 2:**
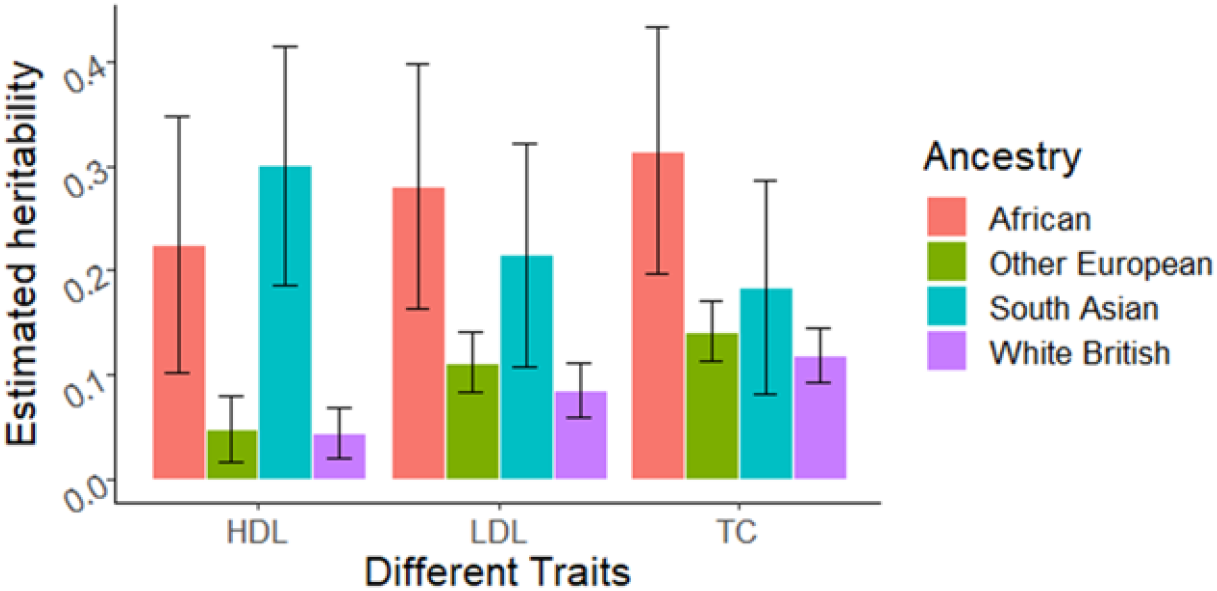
Estimated SNP-based heritability across ancestries for cholesterol traits. The main bars indicate SNP-based heritability estimates, and the error bars indicate 95% confidence intervals. TC= Total-cholesterol, HDL= high-density lipoprotein cholesterol, LDL= low-density lipoprotein cholesterol.

**Figure 3:**
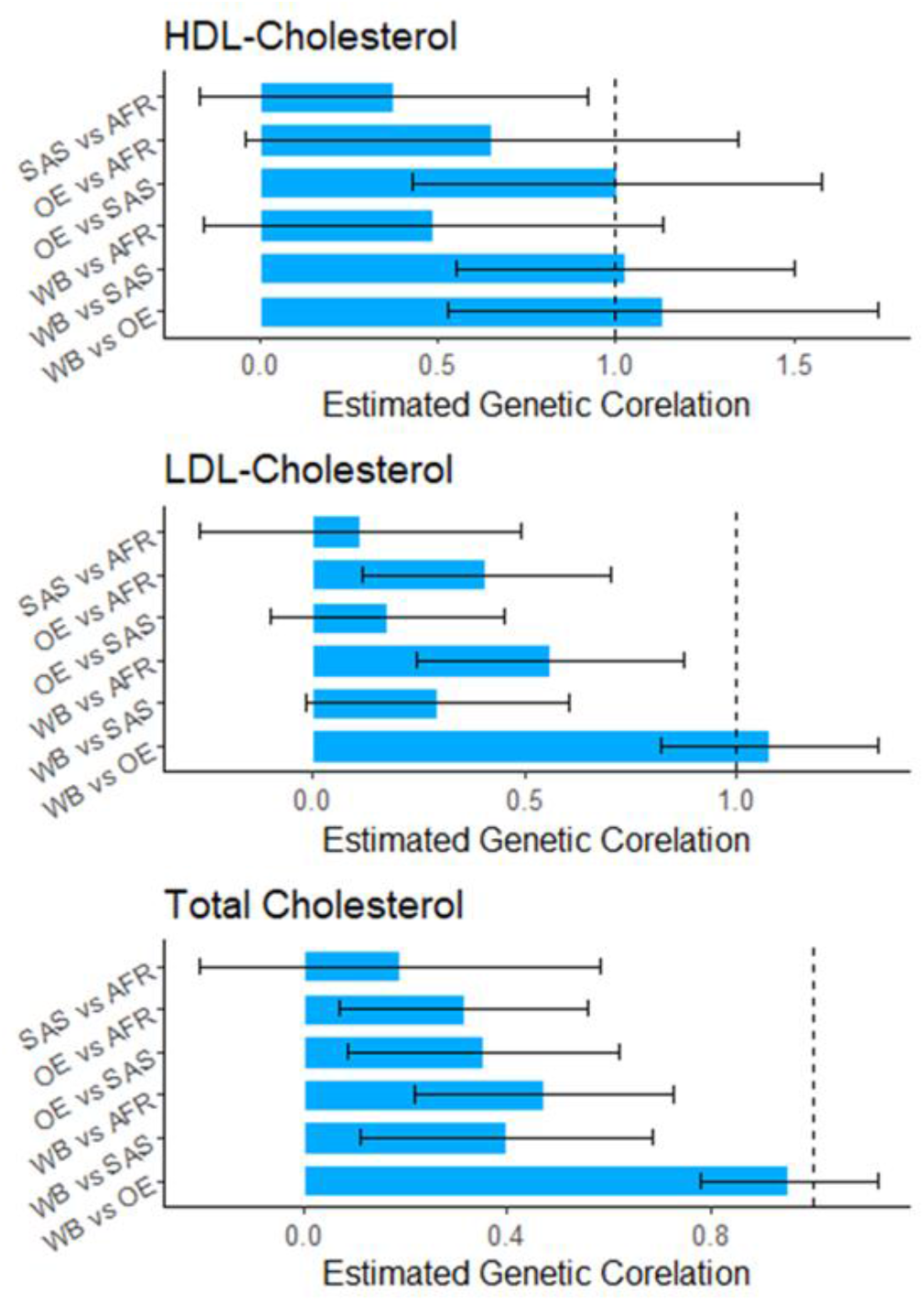
Estimated cross-ancestry genetic correlations. The main bars indicate estimated cross-ancestry genetic correlations, and the error bars indicate 95% confidence intervals of the estimates. WB = White British, OE = Other European, SAS = South Asian, AFR = African.

### Genomic partitioning of cross-ancestry genetic covariance using concordant and discordant SNPs between two diverse ancestries

Some genes are functionally homogeneous across ancestries while the other genes may not be^36, 42, 43^. It can be hypothesised that the functionally homogenous genes are enriched in the regulatory regions, and they contribute more to phenotypic variation within and between ancestries, compared to the other genes. We obtained a set of concordant SNPs (a proxy of functionally homogenous genes) for total-, HDL-, and LDL-cholesterols, by comparing the direction of SNP effects between two diverse ancestries, using the GWAS summary statistics of UK Biobank and Biobank Japan. For the UK Biobank GWAS, we used 258,792 white British individuals who are not overlapping with anyone in the 4 ancestry groups used in our study (white British, other European, South Asian, and African). For the Biobank Japan, we used GWAS summary statistics that are publicly available. In this concordance/discordance analysis, we considered the same HapMap3 SNPs used in the genetic correlation analyses above. The numbers of concordant and discordant SNPs for each pair of ancestries are presented in **Supplementary Table 4**.

First, we quantified if the concordance SNPs are more frequently found in the regulatory or genic region, compared to the other genomic regions for total-cholesterol. **Figure 4** shows that the number of concordant SNPs in the regulatory region is significantly higher than the non-regulatory region (OR= 1.09, *p*-value=2.2e-26 for *p*-value ≤ 1; OR= 1.21, *p*-value=9.2e-16 for *p*-value ≤ 0.05; OR= 1.18, *p*-value=1.6e-06 for *p*-value ≤ 0.01). When selecting SNPs with a genome-wide association (GWA) *p*-value > 0.05 or 0.01, the odds ratio increases (**Figure 4**). Similarly, the number of concordant SNPs in the genic region is significantly higher than the non-genic region (OR= 1.03, *p*-value=1.8e-16 for *p*-value ≤ 1; OR= 1.17, *p*-value=1.6e-32 for *p*-value ≤ 0.05; OR= 1.18, *p*-value=1.8e-06 for *p*-value ≤ 0.01) (**Figure 4**). Similar results were observed when using the HDL- and LDL-cholesterol traits (**Supplementary Figure 1 and 2**).

**Figure 4:**
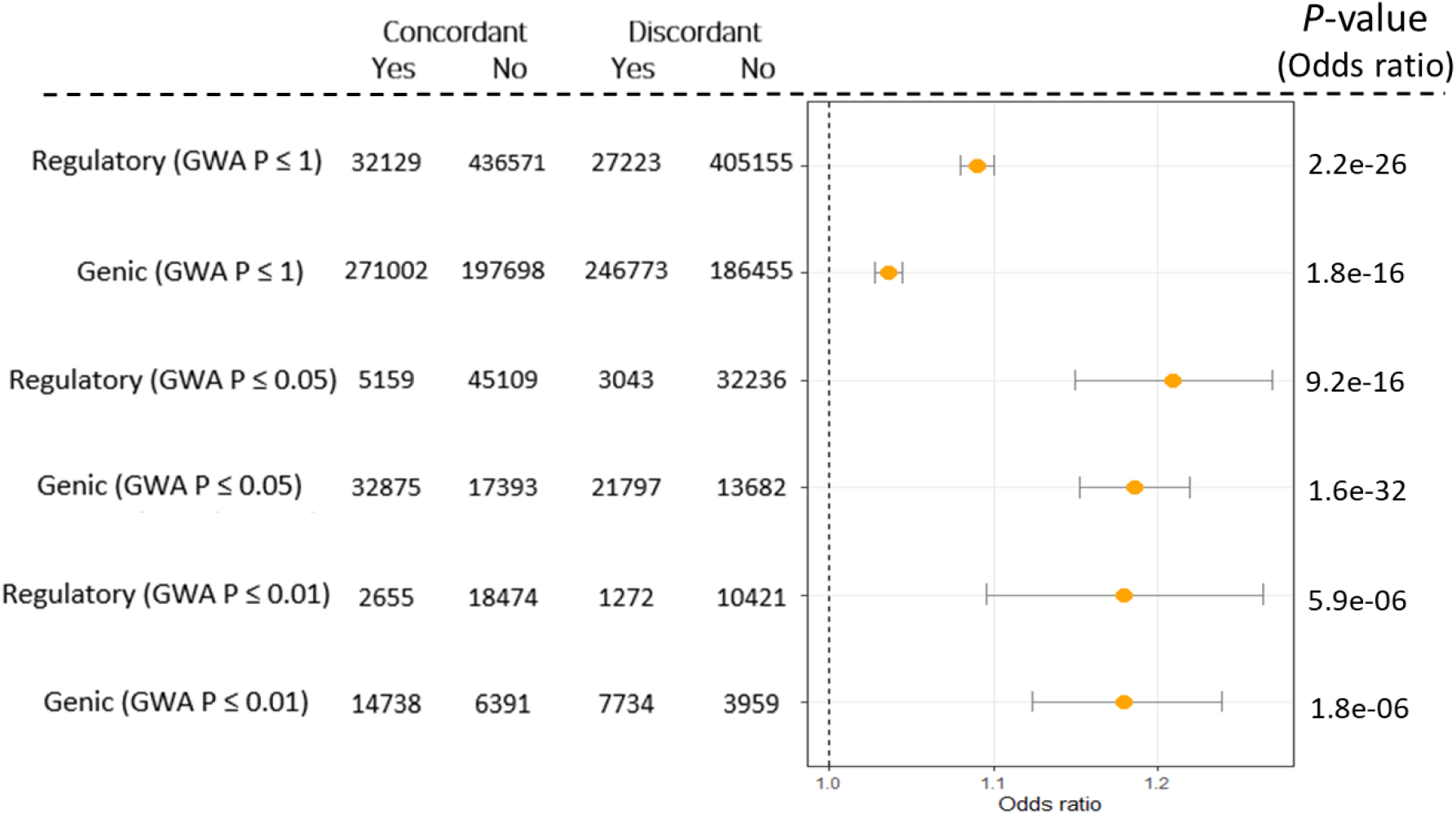
A forest plot with odds ratios indicating that concordant SNPs are more frequently found in the regulatory or genic region. This analysis is for total-cholesterol phenotypes. Error bar represents 95% confidence intervals. The *p*-value of odds ratio indicates that the odds ratio is significantly different from 1. For regulatory or genic region, a genome-wide association (GWA) *p*-value threshold ≤1, 0.05 or 0.01 was used to select a set of concordant and discordant SNPs using UK Biobank GWAS summary statistics for total-cholesterol.

Subsequently, we partitioned genetic covariance components attributed to the two sets of genomic regions (concordant vs. discordant SNPs). We estimated two genomic relationship matrixes (GRM), using the sets of concordant and discordant SNPs, which were simultaneously fitted in a bivariate multiple random-effects model. When considering the set of concordant SNPs, the estimated genetic covariances between other European (OE) vs. south Asian (SAS), white British (WB) vs. African (AFR) and WB vs. OE were significantly higher than the expectation (the proportion of the concordant SNPs) for total-cholesterol (**Figure 5**). On the other hand, the estimated genetic covariances for these pairs of ancestries were significantly lower than the expectation when using discordant SNPs (**Figure 5**). For HDL-cholesterol (**Supplementary Figure 3**) and LDL-cholesterol (**Supplementary Figure 4**), a similar result was observed that the estimated genetic covariances between OE vs. SAS, WB vs. OE and WB vs. SAS were significantly deviated from the expectation. When using SNPs with genome-wide association *p*-values < 0.05 or 0.01 (**Supplementary Table 4**), the estimated genetic covariances due to concordant and discordant SNPs were more significantly deviated from the expectation in general (**Figure 5, Supplementary Figure 3 and 4**). It is also noted that the estimated genetic covariances for the set of discordant SNPs were not higher than zero (Figure 5), implying that the genetic heterogeneity of cholesterol traits across ancestry might be mostly due to the set of discordant SNPs. This also shows that the set of concordant SNPs may be useful in cross-ancestry polygenic risk predictions. The results are similar when genome-wide association p-values from BBJ are used (**Supplementary Figure 5**).

**Figure 5:**
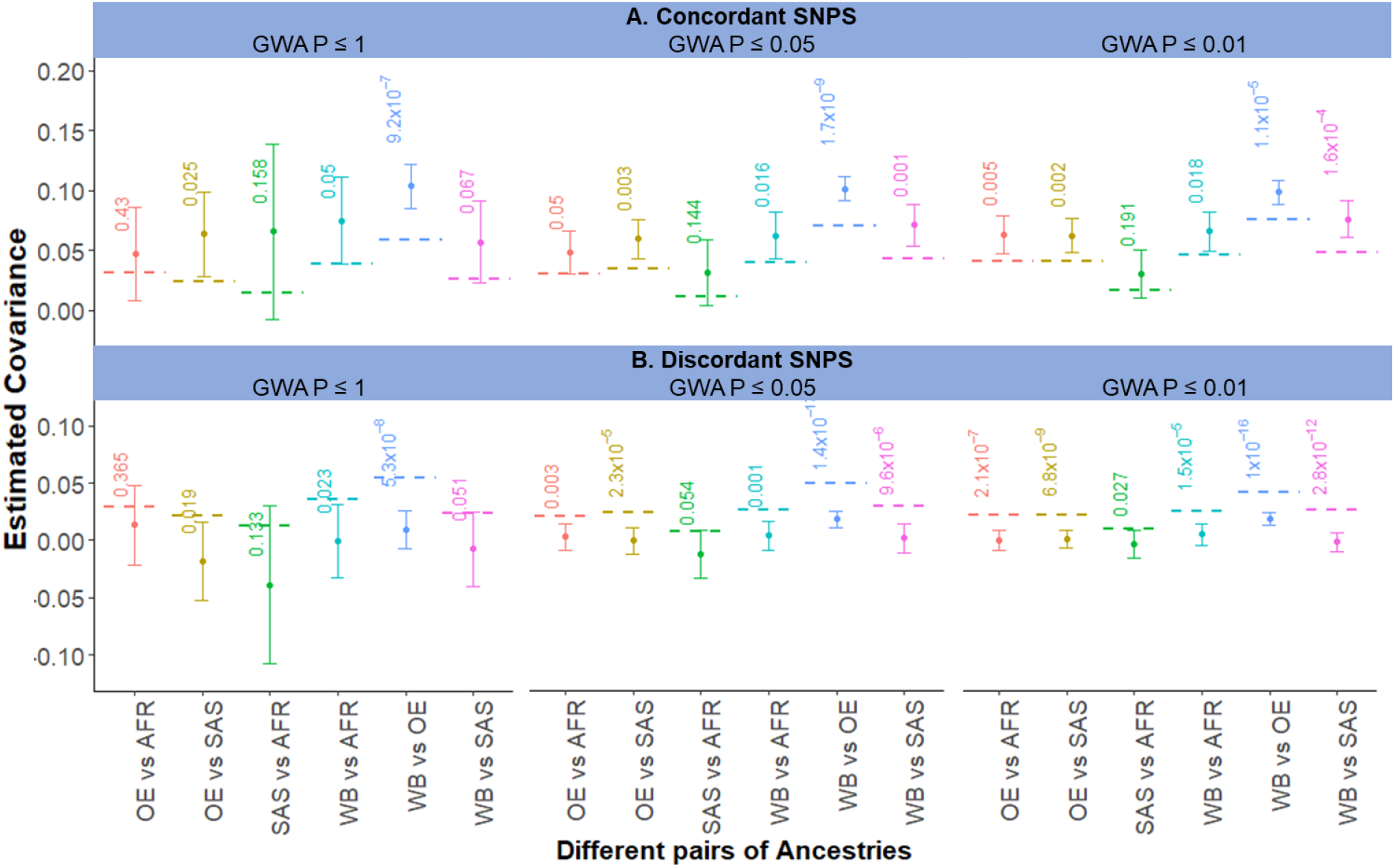
Estimated genetic covariances for concordant and discordant SNPs for total-cholesterol. Concordant and discordant SNPs were derived from the comparison of SNP effects between two independent GWAS datasets of UK Biobank and BBJ. In this concordant or discordant analysis, a set of SNPs with genome-wide association (GWA) *p*-values < 1, 0.05 or 0.01 was used, where the GWA p-values were from UK Biobank GWAS for total-cholesterol. The main bars represent estimated cross-ancestry genetic covariance using the set of genome-wide SNPs, and the error bars indicate 95% confidence intervals. The horizontal dashed line indicates the expected genetic covariance, assuming all SNPs contribute equally to the genetic covariance, i.e., the expected genetic covariance = the estimated total genetic covariance × the proportion of number of concordant SNPs, where the estimated total genetic covariance is based on all the SNPs including both concordant and discordant SNPs. The value with each bar indicates a *p*-value testing the null hypothesis that the estimated genetic covariance is not significantly different from the expectation. WB = White British, OE = Other European, SAS = South Asian, AFR = African.

We further investigated the impact of concordant SNPs in a cross-ancestry polygenic risk prediction. We used the UKBB discovery dataset, which is independent from the four target datasets including white British, other European, South Asian, and African ancestries, to estimate SNP effects and obtain GWAS summary statistics for cholesterol traits. Using the GWAS summary statistics, we constructed polygenic risk scores for the individuals in the target datasets. The predictive ability (*R*^2^) of polygenic risk scores for total-cholesterol is significantly higher when using the set of concordant SNPs than when using the set of discordant SNPs for both within- and cross-ancestry predictions (**Figure 6, Supplementary Figure 4**) (*p*-values for the difference between concordant and discordant PRS SNPs are 3.8e-33, 2.2e-25, 1.3e-04 and 5.3e-04 for white British, other European, South Asian and African, respectively). Although not significant, *R*^2^ is slightly higher when using the set of concordant SNPs, compared to when using the total set of SNPs (**Figure 6**), suggesting that including discordant SNPs may have adverse effects on the cross-ancestry risk predictions. When accounting for the proportion of concordant SNPs, a similar result was observed in that concordant SNPs performed better that discordant SNPs in within- and cross-ancestry risk predictions (**Supplementary Figure 5**). A similar finding was observed when using BBJ discovery GWAS summary statistics, i.e., the cross-ancestry prediction accuracy of the concordant SNPs significantly higher than the discordant SNPs (**Supplementary Figure 4**). Interestingly, the concordant SNPs performs notably better than the total set of SNPs when predicting white British, other European and south Asian ancestries (**Supplementary Figure 4**). Results are invariant when considering LDL- and HDL-cholesterol (Supplementary Figures 6-7).

**Figure 6:**
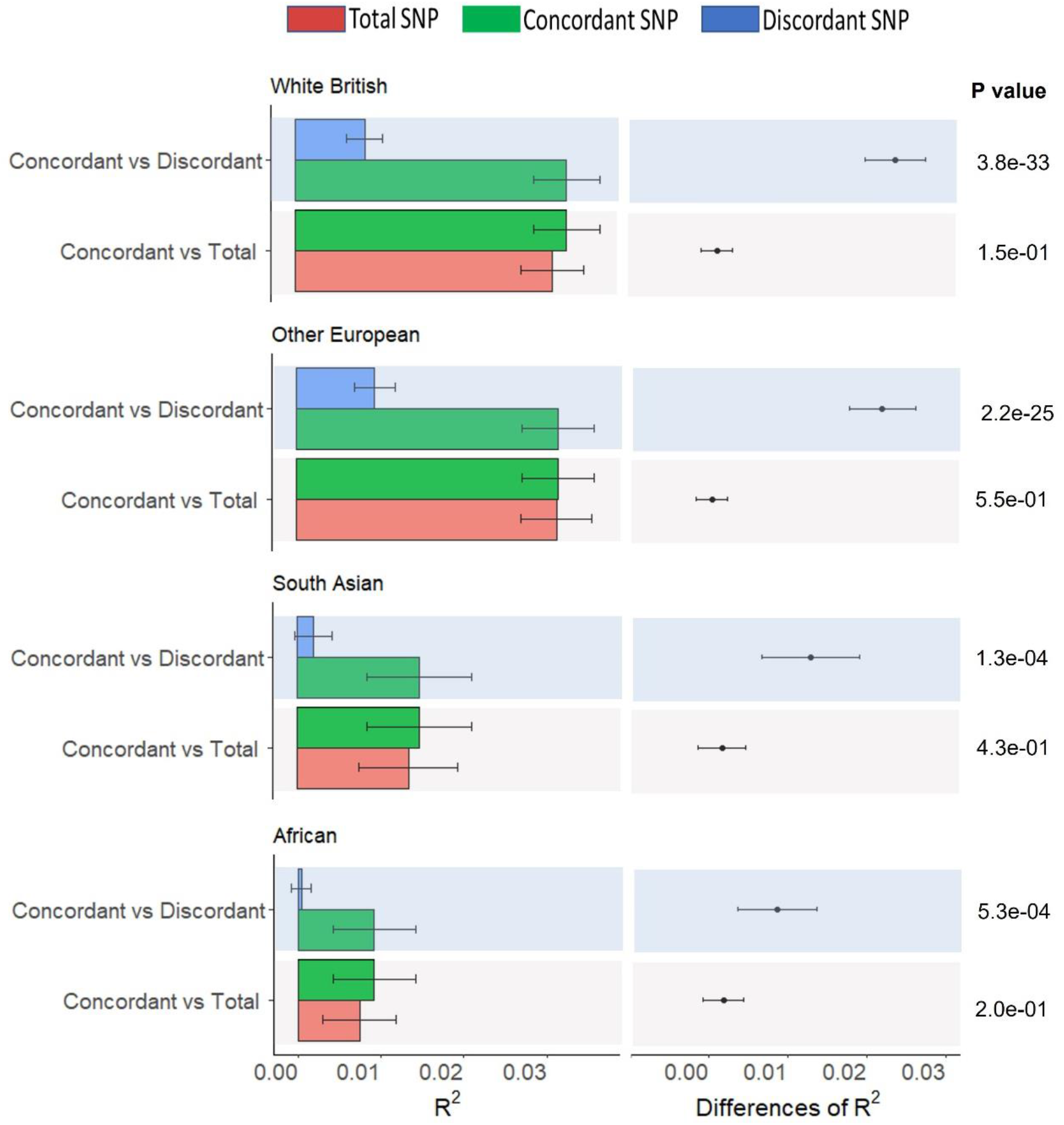
The predictive ability (*R*^2^) of polygenic risk scores for total-cholesterol when using the set of concordant, discordant, or total SNPs for cross-ancestry risk predictions. UK Biobank GWAS was used as the discovery dataset (n= 258,792), while target datasets were other European (n=26,457), south Asian (n=6,199) and African ancestry (n=6,179). **Left panels:** The main bars represent *R*^2^ values and error bars correspond to 95% confidence interval. **Right panels:** Dot points represent the differences between *R*^2^ values, error bars correspond to 95% confidence intervals of the differences, and *p*-values indicate that the differences of *R*^2^ are significantly different from zero (null hypothesis). P-values was estimated using an R-package (r2redux)^45^ based on Wald’s test statistics.

For HDL cholesterol, it is notable that the accuracy of cross-ancestry prediction can be higher than within-ancestry prediction (e.g., South Asian vs. White British in Supplementary Figure 6). We further confirmed this result with a clump-and-threshold (C + T) based PRS method (PRSice)^**44**^ and compared the significance of difference (Supplementary Figure 8). It shows that PGS generated from White British GWAS provides a significantly higher predictive accuracy for South Asian (p-value = 6.67e-16) and African ancestry groups (p-value = 7.35e-04), compared to White British. This may have an important implication in genomic medicine for underrepresented non-European populations.

## Discussion

Cholesterol is an essential structural component of the cell membrane, which is necessary for the body to function^1, 2^. However, the risk of CVD is associated with a high level of cholesterol that can be determined by genetic risk factors^4, 46, 47^. Although the genetic study of cholesterol has been conducted, it is not clear how genetic effects on cholesterol vary across different ancestries. In this study, we explicitly estimated cross-ancestry genetic correlations to investigate the shared genetic architecture across ancestries for cholesterol. Importantly, we appropriately accounted for the relationship between allele frequency and per-allele effect size by modelling the ancestry-specific scale factor for cholesterol, which can provide more reliable estimates^31^.

The reliable estimation of cross-ancestry genetic correlation allows us to understand the shared genetic architecture across ancestries, providing crucial information when for various downstream analyses of complex traits such as cross-ancestry GWAS and cross-ancestry polygenic risk score prediction. Moreover, this may inform best practices for cross-ancestry meta-analysis, multi-ancestry disease mapping, and the transferability of epidemiological findings. Our analysis shows that in general, total- and LDL-cholesterol are both genetically heterogeneous across ancestries, whereas HDL-cholesterol is not^48^. This finding has important implications for the power of cross-ancestry GWASs and cross-ancestry polygenic risk score prediction, which for HDL-cholesterol may be much higher than that for total- and LDL-cholesterols (**Supplementary Figure 6**).

To identify genetic variants that contribute to the genetic heterogeneity, we investigated concordant and discordant SNP sets that were identified by comparing the direction of SNP effects between UK Biobank and Biobank Japan GWAS summary statistics, noting that the two datasets are independent from the four target ancestry groups used in this study. The concordant SNPs may be associated with genes that are functionally homogeneous across ancestries^49^, and we show in this study that the concordant SNPs are more often located in the regulatory or genic regions, compared to other genomic regions. We also show that such strong genetic heterogeneity across ancestries for cholesterol can be attributed to the discordant SNPs, but not to the concordant SNPs. We provide evidence that the set of concordant SNPs can be useful in the cross-ancestry polygenic risk predictions, which may improve the transferability of polygenic risk scores to clinical practice^16, 50, 51^.

There are a number of limitations in this study. For determining optimal *α*, we did not consider the relationship between LD and per-allele effect sizes, i.e., as in LDAK-thin model^32^ that requires a substantial reduction of the number of SNPs. We also acknowledge that the conclusions from cross-ancestry analyses (cross-ancestry correlation and genomic prediction) in this study are restricted to common variants (MAF ≥ 0.01) and HapMap3 SNPs only; as these are robust and reliable for dissecting cross-ancestry genetic architecture^52, 53^. A moderate sample size (limited power of the data) was used to estimate optimal scale factors (*α*) for south Asian and African populations. Therefore, the genetic heterogeneity needs to be explored with larger sample size. The concordant SNPs were identified by comparing the direction of SNP effects between white British (UKBB) and East Asian (BBJ) populations, because adequate data was not available from other ancestries. When public genomic databases have sufficient resources across ancestries, we can have a finer set of concordant SNPs by comparing SNP effects across various ancestries.

In conclusion, there is a significant genetic heterogeneity between ancestries for total- and LDL-cholesterol, which is mostly driven by the set of discordant SNPs. Interestingly, the concordant SNPs are more frequently found in the regulatory region as annotated by an independent study^54^, and restricting to concordant SNPs can provide better accuracy for cross-ancestry polygenic prediction for cholesterol. Our findings contribute to knowledge about the genetic architecture of cholesterol that is shared across ancestries. The proposed cross-ancestry polygenic prediction can be potentially useful in clinical practice. Our analysis protocol can be extended to a wide range of other complex traits and diseases.

## Methods

### Ethical statement

We used publicly available from the UK Biobank (https://www.ukbiobank.ac.uk/). Science protocol and operational procedures for the UK Biobank have been reviewed and approved by the North-West Multi-Centre Research Ethics Committee (MREC), National Information Governance Board for Health & Social Care (NIGB), and Community Health Index Advisory Group (CHIAG). The UK Biobank has obtained consent from all participants. The access of the UK Biobank data was approved under the reference number 14575 (“Whole genome approaches for dissecting (shared) genetic architecture and individual risk prediction of complex traits in human populations”). Publicly available GWAS summary statistics of Biobank Japan (BBJ) were used, following BBJ’s guidelines (http://jenger.riken.jp/en/). The research ethics approval of this study has been obtained from the University of South Australia Human Research Ethics Committee.

### Participants and stratification of ancestries

Data from the UK Biobank contains 501,748 participants recruited between 2006 and 2010^55^. The participants were recruited from 22 assessment centres in England, Wales, and Scotland, ranging in age from 37 to 73 years old^56^. All the phenotypic data for cholesterol traits under this study are derived from baseline survey. Principal component analysis was applied to the UK Biobank individuals to stratify participants^57^ into four different ancestries following previous approach^31^.

### Genotypic data and quality control

We used the second release of the UK biobank (https://www.ukbiobank.ac.uk/) genotype data comprising 488,377 individuals and 92,693,895 imputed autosomal SNPs. The individuals were genotyped by Affymetrix UK BiLEVE Axiom array and Affymetrix UK Biobank Axiom® array. Combination of UK10K and Haplotype Reference Consortium (HRC) data were considered as the reference dataset for the imputation of the UK Biobank genotypic dataset^58^. In this analysis, to dissect the genetic architecture of disease and complex traits, we retained only HapMap3 SNPs in this analysis^52^, which are also considered robust and reliable for estimating heritability, genetic correlation^52, 59^. Stringent quality control (QC) procedure was applied to each ancestry to select high quality individuals and high-quality SNPs. SNPs QC criteria include, SNPs excluded with an INFO score (used to indicate the quality of genotype imputation) <0.6^60-62^, call rate <0.95, a MAF <0.01 and a Hardy–Weinberg equilibrium *p*-value <10^−4^. We also exclude population outliers (individuals outside ±6SD) and related individuals (--rel-cutoff 0.05) using PLINK^63^.

Individual level QC criteria include samples with genotype missing rate >0.05, gender mismatch (reported gender does not fit with the genetically assigned sex determined from gene data), poor genotype quality or a sex chromosome aneuploidy was excluded from the main analyses. For the ease of computation, we reduced the number individuals in white British ancestry. The total number of individuals and total number of SNPs after QC shown in **Supplementary Table 1**. The number of common SNPs across different pairs of ancestries presented in **Supplementary Table 2** and the number of common SNP for each genomic region (genomic partitioning) between ancestries presented in **Supplementary Table 3**.

### Functional annotation of the genome

The common SNPs between populations were partitioned into genomic region using genomic annotation reported by Gusev et. al.^54^, where they partitioned the genome into coding, UTR, promoter, intron, DHS and intergenic regions. For the genomic partitioning analysis, we include promoter, coding, UTR, and DHS regions as regulatory regions^64-66^, and introns (an integral part of a gene)^67, 68^ and the intergenic regions as non-genic regions. We also partitioned the whole genome into two predefined functional categories as genic (includes SNPs from promoter, coding, untranslated, intron and DHS region) and non-genic regions (intergenic region).

### Concordant and discordant SNP annotation

To identify concordant and discordant SNPs we compared SNP effects between two independent GWAS datasets of white British from UK Biobank and Biobank Japan (BBJ). The BBJ summary statistics data are publicly available (http://jenger.riken.jp/en/result). We excluded SNPs that were ambiguous or had a strand issue. After excluding these SNPs, there were 4,113,630 SNPs that are common between UKBB and BBJ. To determine concordant and discordant SNPs, we compared the direction of SNP effects between white British from UKBB and BBJ. We used only HapMap3 SNPs from 4,113,630 SNPs for concordant and discordant analysis across different ancestry pairs (**Supplementary Table 4**).

There were four possible combinations of direction of SNP effects (beta):

(+beta, +beta) if the SNP effects are positive in both GWAS.

(+beta, -beta) if the SNP effects are positive and negative in the UKBB and BBJ GWAS.

(-beta, +beta) if the SNP effects are negative and positive in the UKBB and BBJ GWAS.

(-beta, -beta) if the SNP effects are negative in both GWAS.

Each SNP should be in one of four possible combinations and belongs to either concordance or discordance. SNPs belonged to ((+beta, +beta) ∪ (-beta, -beta) were considered concordant, otherwise discordant, i.e. (+beta, -beta) ∪ (-beta, +beta).

### Data analysis

#### Phenotypic adjustment of main traits

Prior to model fitting, all cholesterol traits were adjusted for demographic variables, the UK biobank assessment centre (as factor), genotype measurement batch (as factor) and population structure measured by the first 10 principal components (PCs)^64, 69^ using linear models in *R*-*software* (4.0.3). Demographic variable includes sex, birth year, education, and Townsend deprivation index (**Supplementary Table 5**). Information of educational qualifications converted to education levels (years) for all the UK Biobank individuals^70^.

### Determining scale factors for GCTA-*α* model

GCTA model assumes all the SNPs has equal contribution to the genetic variance (has no LD weights), whereas LDAK-thin model^32^ explicitly considers LD among SNPs. The previously recommended and widely used α are -0.50 and -0.125 for GCTA model^71^ and LDAK-thin model^32^, respectively. Here we have used 13 different values of *α* (between -1 and 0.5) following GCTA model (termed as GCTA-*α* model)^31^. In order to perform a cross-ancestry genetic correlation analysis of cholesterol traits (total cholesterol, HDL cholesterol, and LDL cholesterol), we determined and used optimal *α* based on GCTA models for each trait and ancestry. We did not consider another widely used LDAK-thin model as it will reduce number of common SNPs between ancestry due to LD-pruning.

### Statistical models

#### Univariate Linear Mixed Model

The univariate Linear Mixed Model (LMM) for can be written as,

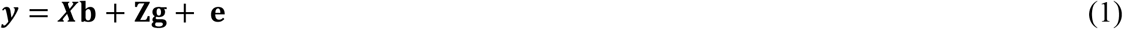

Where **y** is the vector of phenotypic observation, ***b*** is the vector of fixed effects, ***g*** is the vector of additive genetic value and ***e*** is the vector of the residuals. The random effects (***g*** and ***e*** are presumed to be distributed normally with mean zero where **X** and **Z** are incidence matrices

Heritability was estimated using the genetic and residual variances obtained from the univariate LMM, which can be expressed as

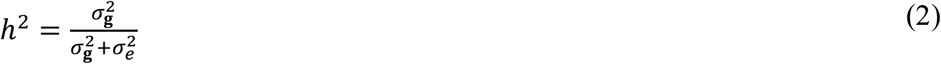

Here, 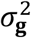 is the genetic variance and 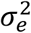 is residual variance. Estimation assumed environmental homogeneity

### Bivariate Linear Mixed Model

The bivariate Linear Mixed Model (LMM) was used to estimate heritability and cross-ancestry genetic correlation using individual level genetic data written as,

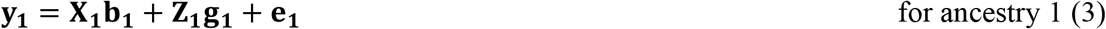

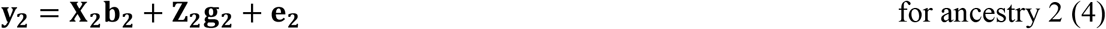

Where ***y***_**1**_ and ***y***_**2**_ are vector of phenotypic observation, **b**_**1**_ and **b**_**2**_ are the vector of fixed effects, **g**_**1**_ **and g**_**2**_ are vector of additive genetic value and **e**_**1**_ and **e**_**2**_ are the vector of residuals. The random effects (**g**_**1**_, **g**_**2**_ and **e**_**1**_, **e**_**2**_) are presumed to be distributed normally with mean zero where **X** and **Z** are incidence matrices i.e. i.e. 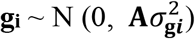 and 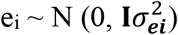.

The variance covariance matrix of observed phenotypes can be written as

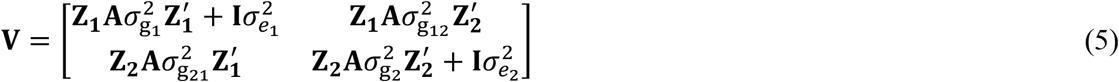

where, **A** is the genomic relationship matrix (GRM)^72-74^, which can be estimated based on the genome-wide SNP information, and **I** is an identity matrix which implicitly assumes across individuals of environmental effects and measurement error. The terms, 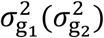 and 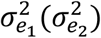 indicate the genetic and residual variance of the trait for the two-ancestry group, and 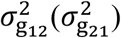 is the genetic covariances between the two ancestry groups. It is noted that there is no parameter to model residual correlation in **V** because there are no multiple phenotypic measures for any individual, i.e., the phenotypes of the first (second) trait are available only for the first (second) ancestry group.

Cross-ancestry genetic correlation between two random genetic effects can be computed either directly as genetic covariance standardized by the square root of the product of the genetic variances of the two random genetic effects (equation 6) or indirectly by the correlation coefficient of SNP effect sizes^38, 75^.

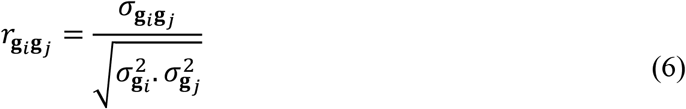

### GREML analysis to estimate heritability and cross-ancestry genetic correlations

Bivariate GREML is the cornerstone method to estimate SNP heritability and cross-ancestry genetic correlation using common SNPs across ancestries. The SNPs frequency, heritability model (relationship between heritability and MAF), and the scale factor (*α*) varied across ancestries^31^. We used a recently proposed approach of estimating GRM^31^ in combined population, that accounts ancestry specific *α* and ancestry specific allele frequencies for estimating heritability and cross-ancestry genetic correlation. Both estimation of GRM and GREML analysis was implemented in *mtg2*^76^.

### Genomic prediction

The polygenic score (PGS) is obtained from by aggregating and quantifying single nucleotide polymorphism (SNP) effects. PGS of an individual (*k*) can be defined as cumulative effect of SNP counts with a standard equation as:

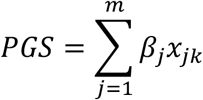

Here, *β*_*j*_ is the SNP effect from discovery GWAS, *m* is the total number of SNPs included in the predictor, *x*_*jk*_ is the number of copies (0,1, or 2) of trait associated SNP *j* in the genotype of individual *k*.

## Data Availability

The genotype and phenotype data of the UK Biobank can be accessed through procedures described on its webpage (https://www.ukbiobank.ac.uk/) and summary statistics of BMI and total-, LDL- and HDL-cholesterol from Biobank Japan (BBJ) can be obtained from its website (http://jenger.riken.jp/en/result)
MTG2, https://sites.google.com/site/honglee0707/mtg2
PLINK2 version can be downloaded from https://www.cog-genomics.org/plink/
r2redux R-package (https://github.com/mommy003/r2redux from GitHub or from CRAN)
The GWAS summary statistics dataset that is generated in this current study and supports the findings have been deposited in the NHGRI-EBI GWAS catalogue with the accession codes GCST90244051, GCST90244052, GCST90244053, GCST90244054, GCST90244055 and GCST90244056; (https://www.ebi.ac.uk/gwas/). GWAS for all SNPs and concordant SNPs for total-, HDL- and LDL-cholesterol can be accessed in following links
GWAS of total cholesterol (all SNP) (https://ftp.ebi.ac.uk/pub/databases/gwas/summary_statistics/GCST90244001-GCST90245000/GCST90244051/)
GWAS of total cholesterol (concordant SNP) (https://ftp.ebi.ac.uk/pub/databases/gwas/summary_statistics/GCST90244001-GCST90245000/GCST90244052/)
GWAS of HDL-cholesterol (all SNP) (https://ftp.ebi.ac.uk/pub/databases/gwas/summary_statistics/GCST90244001-GCST90245000/GCST90244053/)
GWAS for HDL-cholesterol (concordant SNP) (https://ftp.ebi.ac.uk/pub/databases/gwas/summary_statistics/GCST90244001-GCST90245000/GCST90244054/)
GWAS for LDL-cholesterol (all SNP) (https://ftp.ebi.ac.uk/pub/databases/gwas/summary_statistics/GCST90244001-GCST90245000/GCST90244055/)
GWAS for LDL-cholesterol (concordant SNP) (https://ftp.ebi.ac.uk/pub/databases/gwas/summary_statistics/GCST90244001-GCST90245000/GCST90244056/)

## Web resources and code availability

The genotype and phenotype data of the UK Biobank can be accessed through procedures described on its webpage (https://www.ukbiobank.ac.uk/) and summary statistics of BMI and total-, LDL- and HDL-cholesterol from Biobank Japan (BBJ) can be obtained from its website (http://jenger.riken.jp/en/result)

MTG2, https://sites.google.com/site/honglee0707/mtg2

PLINK2 version can be downloaded from https://www.cog-genomics.org/plink/

*r2redux* R-package (https://github.com/mommy003/r2redux from GitHub or from CRAN)

The GWAS summary statistics dataset that is generated in this current study and supports the findings have been deposited in the NHGRI-EBI GWAS catalogue with the accession codes GCST90244051, GCST90244052, GCST90244053, GCST90244054, GCST90244055 and GCST90244056; (https://www.ebi.ac.uk/gwas/). GWAS for all SNPs and concordant SNPs for total-, HDL- and LDL-cholesterol can be accessed in following links

GWAS of total cholesterol (all SNP) (https://ftp.ebi.ac.uk/pub/databases/gwas/summary_statistics/GCST90244001-GCST90245000/GCST90244051/)

GWAS of total cholesterol (concordant SNP) (https://ftp.ebi.ac.uk/pub/databases/gwas/summary_statistics/GCST90244001-GCST90245000/GCST90244052/)

GWAS of HDL-cholesterol (all SNP) (https://ftp.ebi.ac.uk/pub/databases/gwas/summary_statistics/GCST90244001-GCST90245000/GCST90244053/)

GWAS for HDL-cholesterol (concordant SNP) (https://ftp.ebi.ac.uk/pub/databases/gwas/summary_statistics/GCST90244001-GCST90245000/GCST90244054/)

GWAS for LDL-cholesterol (all SNP) (https://ftp.ebi.ac.uk/pub/databases/gwas/summary_statistics/GCST90244001-GCST90245000/GCST90244055/)

GWAS for LDL-cholesterol (concordant SNP) (https://ftp.ebi.ac.uk/pub/databases/gwas/summary_statistics/GCST90244001-GCST90245000/GCST90244056/)

## Acknowledgements

This research is supported by the Australian Research Council (DP190100766). We thank the staff and participants of the UK Biobank and Biobank Japan for their important contributions. Our reference number approved by UK Biobank is 14575. The analyses were performed using computational resources provided by the Australian Government through Gadi under the National Computational Merit Allocation Scheme (NCMAS), and HPCs (Statgen server) managed by UniSA IT.

