## Supplementary information for "Cross-ancestry genetic architecture and prediction for cholesterol traits"

**Supplementary Table 1: The number of individuals and SNPs in different ancestry groups in the analyses of cross-ancestry genetic correlations**

| **Ancestry** | **Initial sample size** | **Individuals after QC** | **SNPs after QC** |
| --- | --- | --- | --- |
| **White British** | 430,301 | 30,000^*^ | 1,154,490 |
| **Other European** | 29,023 | 26,457 | 1,148,504 |
| **South Asian** | 7,449 | 6,199 | 939,512 |
| **African** | 7,647 | 6,179 | 729,534 |

^*^We randomly selected 30000 individuals from the total QCed white British ancestry individuals for estimating cross-ancestry genetic correlations because it is computationally feasible in multiple analyses comparing other ancestries.

**Supplementary Table 2:** The number of common SNPs (after QC) between ancestries

| **Ancestry** | **White British** | **Other European** | **South Asian** | **African** |
| --- | --- | --- | --- | --- |
| **White British** |  | 1,138,117 | 904,421 | 632,906 |
| **Other European** | - |  | 909,472 | 635,763 |
| **South Asian** | - | - |  | 609,243 |
| **African** | - | - | - |  |

**Supplementary Table 3: Genomic partitioning based on commons SNPs (percentage) across different genomic regions across ancestries**

|  | **White British** | **Other European** | **South Asian** | **African** |
| --- | --- | --- | --- | --- |
| **White British** |  | Regulatory= 73,942 (6.54)  Intron= 314,292 (27.79)  Intergenic= 483,867 (42.79)  DHS= 258,776 (22.88) | Regulatory= 63,273 (7.05)  Intron= 253,543 (28.24)  Intergenic= 378,876 (42.20)  DHS= 202,068 (22.51) | Regulatory= 49,462 (7.89)  Intron= 180,803 (28.84)  Intergenic= 256,977 (40.98)  DHS= 139,751 (22.29) |
| **Other European** |  |  | Regulatory= 63,767 (7.06)  Intron= 254,826 (28.21)  Intergenic= 381,539 (42.24)  DHS= 203,043 (22.48) | Regulatory= 49,800 (7.90)  Intron= 181,565 (28.81)  Intergenic= 258,620 (41.03)  DHS= 140,262 (22.26) |
| **South Asian** |  |  |  | Regulatory= 48,763 (8.07)  Intron= 174,889 (28.98)  Intergenic= 246,225 (40.79)  DHS= 133,702 (22.15) |
| **African** |  |  |  |  |

Regulatory regions include coding regions, promoter regions and untranslated regions (UTR). Genic regions include SNPs from regulatory regions, introns and DHS regions, while intergenic regions considered as non-genic regions.

**Supplementary Table 4:** The number and percentage of common concordant and discordant SNPs from HapMap3 SNPs for each pair of ancestries across traits

| **P value Threshold** | **Traits** | **Ancestries** | **N concordant SNPs** | **N discordant SNPs** |
| --- | --- | --- | --- | --- |
| **P ≤ 1** | **Total-cholesterol** | White British vs Other European | 462,719 (51.99%) | 427,313 (48.01%) |
|  |  | White British vs South Asian | 366,856 (52.14%) | 336,710 (47.86%) |
|  |  | White British vs African | 259,545 (52.44%) | 235,439 (47.66%) |
|  |  | Other European vs South Asian | 366,738 (52.15%) | 336,501 (47.85%) |
|  |  | Other European vs African | 259,340 (52.46%) | 235,059 (47.64%) |
|  |  | South Asian vs African | 245,349 (52.50%) | 221,963 (47.50%) |
|  | **HDL-cholesterol** | White British vs Other European | 462,579 (51.97%) | 427,453 (48.03%) |
|  |  | White British vs South Asian | 366202 (52.05%) | 337,364 (47.95%) |
|  |  | White British vs African | 258,200 (52.16%) | 236,784 47.84%) |
|  |  | Other European vs South Asian | 366,109 (52.06%) | 337,130 (47.94%) |
|  |  | Other European vs African | 257,981 (52.18%) | 236,418 (47.82%) |
|  |  | South Asian vs African | 243,818 (52.17%) | 223,494 (47.83%) |
|  | **LDL-cholesterol** | White British vs Other European | 456,663 (51.31%) | 433,369 (48.69%) |
|  |  | White British vs South Asian | 361,860 (51.43%) | 341,706 (48.57%) |
|  |  | White British vs African | 255,268 (51.57%) | 239,716 (48.43%) |
|  |  | Other European vs South Asian | 361,753 (51.44%) | 341,486 (48.46%) |
|  |  | Other European vs African | 254,973 (51.57%) | 239,426 (48.43%) |
|  |  | South Asian vs African | 241,102 (51.59%) | 226,210 (48.41%) |
| **P ≤ 0.05** | **Total-cholesterol** | White British vs Other European | 49,707 (58.79%) | 34,847 (41.21%) |
|  |  | White British vs South Asian | 29,349 (58.90%) | 20,479 (41.10%) |
|  |  | White British vs African | 21,224 (59.02%) | 14,738 (40.98%) |
|  |  | Other European vs South Asian | 29,291 (58.89%) | 20,447 (41.11%) |
|  |  | Other European vs African | 21,182 (59.02%) | 14,705 (40.98%) |
|  |  | South Asian vs African | 20,189 (59.14%) | 13,946 (40.86%) |
|  | **HDL-cholesterol** | White British vs Other European | 40,861 (57.56%) | 30,132 (42.44%) |
|  |  | White British vs South Asian | 33,369 (58.06%) | 24,104 (41.94%) |
|  |  | White British vs African | 24,241 (58.15%) | 17,445 (41.85%) |
|  |  | Other European vs South Asian | 33,317 (58.07%) | 24,060 (41.93%) |
|  |  | Other European vs African | 24,177 (58.19%) | 17,367 (41.81%) |
|  |  | South Asian vs African | 23,059 (58.27%) | 16,513 (41.73%) |
|  | **LDL-cholesterol** | White British vs Other European | 43,539 (56.04%) | 34,149 (43.96%) |
|  |  | White British vs South Asian | 35,872 (56.56%) | 27,546 (43.44%) |
|  |  | White British vs African | 26,620 (56.98%) | 20,098 (43.02%) |
|  |  | Other European vs South Asian | 35,744 (56.50%) | 27,519 (43.50%) |
|  |  | Other European vs African | 26,474 (56.90%) | 20,053 (43.10%) |
|  |  | South Asian vs African | 25,278 (56.95%) | 19,106 (43.05%) |
| **P ≤ 0.01** | **Total-cholesterol** | White British vs Other European | 10,965 (63.80%) | 6,222 (36.20%) |
|  |  | White British vs South Asian | 9,068 (64.33%) | 5,028 (35.67%) |
|  |  | White British vs African | 6,719 (64.40%) | 3,708 (35.60%) |
|  |  | Other European vs South Asian | 9,050 (64.39%) | 5,006 (35.61%) |
|  |  | Other European vs African | 6,708 (64.54%) | 3,685 (35.46%) |
|  |  | South Asian vs African | 64,03 (64.72%) | 3,490 (35.28%) |
|  | **HDL-cholesterol** | White British vs Other European | 14,166 (62.23%) | 8,598 (37.77%) |
|  |  | White British vs South Asian | 11,890 (63.12%) | 6,947 (36.88%) |
|  |  | White British vs African | 8,805 (63.58%) | 5,044 (36.42%) |
|  |  | Other European vs South Asian | 11,881 (63.14%) | 6,937 (36.86%) |
|  |  | Other European vs African | 8,792 (63.60%) | 5,031 (36.40%) |
|  |  | South Asian vs African | 8,467 (63.90%) | 4,783 (36.10%) |
|  | **LDL-cholesterol** | White British vs Other European | 16,671 (59.91%) | 11,157 (40.09%) |
|  |  | White British vs South Asian | 14,197 (60.64%) | 9,215 39.36(%) |
|  |  | White British vs African | 10,858 (61.02%) | 6,936 (38.98%) |
|  |  | Other European vs South Asian | 14,083 (60.49%) | 9,199 (39.51%) |
|  |  | Other European vs African | 10,741 (60.87%) | 6,906 (39.13%) |
|  |  | South Asian vs African | 10,400 (60.95%) | 6,662 (39.05%) |

**Supplementary Table 5: The list of covariates used for adjustment of the cholesterol traits**

| **Variable** | **Data-field in UK Biobank** | **Value Type** |
| --- | --- | --- |
| Age at recruitment (age) | 21022 | Integral, years (37-73) |
| Sex (sex) | 31 | Single (male, female) |
| Educational Qualification (EDU)* | 6138 | Categorial (multiple) |
| Birth year (BY) | 34 | Integer, years (1934 to 1971) |
| Social economic status (Townsend deprivation index) (TDI) | 189 | Continuous, (-6.25826 – 11.0013) |
| UK Biobank assessment centre (centre) | 54 | As factor Categorical (single) (26) |
| Genotype measurement batch (geno_batch) | 22000 | As factor (Categorical) |
| Principle component (PC1-PC10) | 22009 | Continuous |

*Information of educational qualifications converted to education levels (years) for all the UK Biobank individuals^1^. From the multiple responses, we consider the highest level of converted education year for each participant for the adjustment of main variable following Shin et al.^2^

**Supplementary Table 6: Determining trait specific scale factor (α) for White British**

| **Log-likelihood** | | | | | | | | | | | | | |
| --- | --- | --- | --- | --- | --- | --- | --- | --- | --- | --- | --- | --- | --- |
| **Traits** | **-1** | **-0.875** | **-0.75** | ***α*= -0.625** | ***α*= -0.50** | ***α*= -0.375** | ***α*= -0.25** | ***α*= -0.125** | ***α*= 0** | ***α*= 0.125** | ***α*= 0.25** | ***α*= 0.375** | ***α*= 0.5** |
| **TC** | -14076.8 | -14067.01 | -14060.43 | -14057.3 | **-14056.52** | -14056.96 | -14057.94 | -14059.11 | -14060.32 | -14061.51 | -14062.63 | -14063.69 | -14064.68 |
| **HDL** | -12964.3 | -12962.46 | -12960.88 | -12959.92 | -12959.5 | -12959.42 | **-12959.14** | -12959.75 | -12959.99 | -12960.23 | -12960.47 | -12960.68 | -12960.87 |
| **LDL** | -14082.6 | -14078.77 | -14076.82 | **-14076.55** | -14077.18 | -14078.11 | -14079.07 | -14079.96 | -14080.76 | -14081.48 | -14082.13 | -14082.71 | -14083.24 |
| **Delta AIC** | | | | | | | | | | | | | |
| **Traits** | **-1** | **-0.875** | **-0.75** | ***α*= -0.625** | ***α*= -0.50** | ***α*= -0.375** | ***α*= -0.25** | ***α*= -0.125** | ***α*= 0** | ***α*= 0.125** | ***α*= 0.25** | ***α*= 0.375** | ***α*= 0.5** |
| **TC** | 40.494 | 20.983 | 7.810 | 1.561 | **0** | 0.881 | 2.837 | 5.183 | 7.607 | 9.972 | 12.223 | 14.342 | 16.326 |
| **HDL** | 10.395 | 6.6402 | 3.486 | 1.562 | 0.717 | 0.567 | **0** | 1.217 | 1.702 | 2.193 | 2.660 | 3.086 | 3.4674 |
| **LDL** | 12.166 | 4.438 | 0.538 | **0** | 1.255 | 3.122 | 5.044 | 6.827 | 8.432 | 9.867 | 11.155 | 12.318 | 13.374 |

TC= Total-cholesterol, HDL= High-density lipoprotein cholesterol, LDL= Low-density lipoprotein cholesterol. Akaike Information Criterion $\left( \mathrm{AIC} \right)=2k-2ln(L)$ where $ln(L)$ is the logarithm of the maximum likelihood from the model and k is the number of model parameters in the model. ΔAIC = AIC – AIC of the best model with the optimal α. The best model is highlighted.

**Supplementary Table 7: Determining trait specific scale factor (α) for the other Europeans**

| **Log-likelihood** | | | | | | | | | | | | | |
| --- | --- | --- | --- | --- | --- | --- | --- | --- | --- | --- | --- | --- | --- |
| **Traits** | **-1** | **-0.875** | **-0.75** | ***α*= -0.625** | ***α*= -0.50** | ***α*= -0.375** | ***α*= -0.25** | ***α*= -0.125** | ***α*= 0** | ***α*= 0.125** | ***α*= 0.25** | ***α*= 0.375** | ***α*= 0.5** |
| **TC** | -12215.17 | -12204.17 | -12196.38 | -12192.23 | -12190.71 | **-12190.71** | -12191.52 | -12192.71 | -12194.06 | -12195.46 | -12196.83 | -12198.15 | -12199.38 |
| **HDL** | -11292.1 | -11291.77 | **-11291.73** | -11291.87 | -11292.04 | -11292.2 | -11292.32 | -11292.42 | -11292.5 | -11292.56 | -11292.61 | -11292.67 | -11292.71 |
| **LDL** | -12226.11 | -12220.06 | -12216.36 | -12214.88 | **-12214.81** | -12215.44 | -12216.39 | -12217.45 | -12218.52 | -12219.56 | -12220.54 | -12221.46 | -12222.29 |
| **Delta AIC** | | | | | | | | | | | | | |
| **Traits** | **-1** | **-0.875** | **-0.75** | ***α*= -0.625** | ***α*= -0.50** | ***α*= -0.375** | ***α*= -0.25** | ***α*= -0.125** | ***α*= 0** | ***α*= 0.125** | ***α*= 0.25** | ***α*= 0.375** | ***α*= 0.5** |
| **TC** | 48.919 | 26.906 | 11.331 | 3.036 | 0.0002 | **0** | 1.604 | 3.983 | 6.692 | 9.487 | 12.236 | 14.864 | 17.336 |
| **HDL** | 0.738 | 0.082 | **0** | 0.268 | 0.621 | 0.935 | 1.182 | 1.373 | 1.525 | 1.651 | 1.762 | 1.864 | 1.961 |
| **LDL** | 22.599 | 10.512 | 3.103 | 0.148 | **0** | 1.27 | 3.164 | 5.283 | 7.431 | 9.510 | 11.473 | 13.297 | 14.975 |

TC= Total-cholesterol, HDL= High-density lipoprotein cholesterol, LDL= Low-density lipoprotein cholesterol. Akaike Information Criterion $\left( \mathrm{AIC} \right)=2k-2ln(L)$ where $ln(L)$ is the logarithm of the maximum likelihood from the model and k is the number of model parameters in the model. ΔAIC = AIC – AIC of the best model with the optimal α. The best model is highlighted.

**Supplementary Table 8: Determining trait specific scale factor (α) for South Asians**

| **Log-likelihood** | | | | | | | | | | | | | |
| --- | --- | --- | --- | --- | --- | --- | --- | --- | --- | --- | --- | --- | --- |
| **Traits** | **-1** | **-0.875** | **-0.75** | ***α*= -0.625** | ***α*= -0.50** | ***α*= -0.375** | ***α*= -0.25** | ***α*= -0.125** | ***α*= 0** | ***α*= 0.125** | ***α*= 0.25** | ***α*= 0.375** | ***α*= 0.5** |
| **TC** | -2779.386 | -2778.078 | -2776.784 | -2775.82 | -2775.216 | -2774.887 | -2774.727 | -2774.664 | **-2774.658** | -2774.685 | -2774.732 | -2774.791 | -2774.856 |
| **HDL** | -2530.547 | -2527.662 | -2524.867 | -2522.76 | -2521.411 | -2520.644 | -2520.242 | -2520.058 | **-2520.004** | -2520.026 | -2520.095 | -2520.192 | -2520.308 |
| **LDL** | -2774.391 | -2772.671 | -2771.031 | -2769.84 | -2769.122 | -2768.745 | -2768.576 | **-2768.528** | -2768.553 | -2768.621 | -2768.717 | -2768.831 | -2768.956 |
| **Delta AIC** | | | | | | | | | | | | | |
| **Traits** | **-1** | **-0.875** | **-0.75** | ***α*= -0.625** | ***α*= -0.50** | ***α*= -0.375** | ***α*= -0.25** | ***α*= -0.125** | ***α*= 0** | ***α*= 0.125** | ***α*= 0.25** | ***α*= 0.375** | ***α*= 0.5** |
| **TC** | 9.457 | 6.839 | 4.252 | 2.314 | 1.115 | 0.458 | 0.137 | 0.012 | **0** | 0.054 | 0.148 | 0.265 | 0.396 |
| **HDL** | 21.088 | 15.316 | 9.726 | 5.503 | 2.816 | 1.281 | 0.477 | 0.109 | **0** | 0.045 | 0.182 | 0.378 | 0.608 |
| **LDL** | 11.725 | 8.285 | 5.005 | 2.619 | 1.187 | 0.433 | 0.095 | **0** | 0.048 | 0.185 | 0.378 | 0.605 | 0.855 |

TC= Total-cholesterol, HDL= High-density lipoprotein cholesterol, LDL= Low-density lipoprotein cholesterol. Akaike Information Criterion $\left( \mathrm{AIC} \right)=2k-2ln(L)$ where $ln(L)$ is the logarithm of the maximum likelihood from the model and k is the number of model parameters in the model. ΔAIC = AIC – AIC of the best model with the optimal α. The best model is highlighted.

**Supplementary Table 9: Determining trait specific scale factor (α) for Africans**

| **Log-likelihood** | | | | | | | | | | | | | |
| --- | --- | --- | --- | --- | --- | --- | --- | --- | --- | --- | --- | --- | --- |
| **Traits** | **-1** | **-0.875** | **-0.75** | ***α*= -0.625** | ***α*= -0.50** | ***α*= -0.375** | ***α*= -0.25** | ***α*= -0.125** | ***α*= 0** | ***α*= 0.125** | ***α*= 0.25** | ***α*= 0.375** | ***α*= 0.5** |
| **TC** | -2779.441 | -2776.842 | -2773.308 | -2769.98 | -2767.838 | -2766.884 | **-2766.68** | -2766.855 | -2767.203 | -2767.623 | -2768.063 | -2768.502 | -2768.928 |
| **HDL** | -2572.669 | -2572.215 | **-2572.183** | -2572.743 | -2573.618 | -2574.413 | -2574.974 | -2575.322 | -2575.525 | -2575.638 | -2575.7 | -2575.732 | -2575.749 |
| **LDL** | -2773.498 | -2771.468 | -2768.729 | -2766.156 | -2764.52 | -2763.815 | **-2763.683** | -2763.83 | -2764.099 | -2764.414 | -2764.741 | -2765.064 | -2765.376 |
| **Delta AIC** | | | | | | | | | | | | | |
| **Traits** | **-1** | **-0.875** | **-0.75** | ***α*= -0.625** | ***α*= -0.50** | ***α*= -0.375** | ***α*= -0.25** | ***α*= -0.125** | ***α*= 0** | ***α*= 0.125** | ***α*= 0.25** | ***α*= 0.375** | ***α*= 0.5** |
| **TC** | 25.523 | 20.326 | 13.257 | 6.601 | 2.316 | 0.409 | **0** | 0.351 | 1.048 | 1.886 | 2.767 | 3.645 | 4.496 |
| **HDL** | 0.972 | 0.064 | **0** | 1.121 | 2.869 | 4.461 | 5.582 | 6.279 | 6.684 | 6.911 | 7.034 | 7.099 | 7.133 |
| **LDL** | 19.631 | 15.571 | 10.092 | 4.947 | 1.674 | 0.264 | **0** | 0.295 | 0.832 | 1.462 | 2.116 | 2.763 | 3.386 |

TC= Total-cholesterol, HDL= High-density lipoprotein cholesterol, LDL= Low-density lipoprotein cholesterol. Akaike Information Criterion $\left( \mathrm{AIC} \right)=2k-2ln(L)$ where $ln(L)$ is the logarithm of the maximum likelihood from the model and k is the number of model parameters in the model. ΔAIC = AIC – AIC of the best model with the optimal α. The best model is highlighted.

**Supplementary Table 10: The estimated cross-ancestry genetic correlation (SE) for total-cholesterol**

|  | **Cohort1/**  **White British**  **(n=28,196)** | **Cohort2/**  **Other European**  **(n=24,487)** | **Cohort3/**  **South Asian**  **(n=5,555)** | **Cohort4/**  **African**  **(n=5,559)** |
| --- | --- | --- | --- | --- |
| **Cohort1/**  **White British** |  | **0.954 (0.087)**  **P= 5.96e-01**  $h_{c}^{2} (WB)$= 0.121 (0.013)  $h_{c}^{2} (OE)$= 0.143 (0.014) | **0.399 (0.143)**  **P= 2.63e-05**  $h_{c}^{2} (WB)$=0.112 (0.012)  $h_{c}^{2} (As)$=0.186 (0.051) | **0.473 (0.127)**  **P= 3.33e-05**  $h_{c}^{2} (WB)$= 0.109 (0.011)  $h_{c}^{2} (Af)$= 0.315 (0.057) |
| **Cohort2/**  **Other European** |  |  | **0.353 (0.133)**  **P=1.14e-06**  $h_{c}^{2} (OE)$= 0.134 (0.014)  $h_{c}^{2} (As)$= 0.191 (0.051) | **0.315 (0.122)**  **P= 1.96e-08**  $h_{c}^{2} (OE)$= 0.127 (0.013)  $h_{c}^{2} (Af)$= 0.316 (0.057) |
| **Cohort3/**  **South Asian** |  |  |  | **0.188 (0.197)**  **P= 3.76e-05**  $h_{c}^{2} (As)$=0.175 (0.046)  $h_{c}^{2} (Af)$= 0.308 (0.056) |
| **Cohort4/**  **African** |  |  |  |  |

P is the *p*-value based on Wald’s test statistics for the null hypothesis of the estimated cross ancestry genetic correlation $r_{g}$=1 (i.e., a two-sided test). $h_{c}^{2}$ is the estimated SNP-based heritability from the bivariate GREML, using SNPs common between two ancestries. WB, OE, SAS, AFR and MA indicates White British, Other European, South Asian, African, and Mixed ancestry cohorts.

**Supplementary Table 11: The estimated cross-ancestry genetic correlation (SE) for LDL-cholesterol**

|  | **Cohort1/**  **White British**  **(n=28,151)** | **Cohort2/**  **Other European**  **(n=24,436)** | **Cohort3/**  **South Asian**  **(n=5,547)** | **Cohort4/**  **African**  **(n=5,547)** |
| --- | --- | --- | --- | --- |
| **Cohort1/**  **White British** |  | **1.084 (0.128)**  **P= 5.12e-01**  $h_{c}^{2} (WB)$= 0.086 (0.013)  $h_{c}^{2} (OE)$= 0.113 (0.014) | **0.296 (0.155)**  **P= 5.57e-06**  $h_{c}^{2} (WB)$= 0.079 (0.012)  $h_{c}^{2} (As)$= 0.215 (0.053) | **0.561 (0.158)**  **P= 5.46e-03**  $h_{c}^{2} (WB)$= 0.084 (0.011)  $h_{c}^{2} (Af)$= 0.284 (0.056) |
| **Cohort2/**  **Other European** |  |  | **0.177 (0.138)**  **P=2.46e-09**  $h_{c}^{2} (OE)$= 0.104 (0.014)  $h_{c}^{2} (As)$= 0.219 (0.053) | **0.409 (0.147)**  **P= 5.81e-05**  $h_{c}^{2} (OE)$= 0.099 (0.013)  $h_{c}^{2} (Af)$= 0.281 (0.056) |
| **Cohort3/**  **South Asian** |  |  |  | **0.110 (0.190)**  **P= 2.81e-06**  $h_{c}^{2} (As)$= 0.209 (0.048)  $h_{c}^{2} (Af)$= 0.277 (0.056) |
| **Cohort4/**  **African** |  |  |  |  |

P is the *p*-value based on Wald’s test statistics for the null hypothesis of the estimated cross ancestry genetic correlation $r_{g}$=1 (i.e., a two-sided test). $h_{c}^{2}$ is the estimated SNP-based heritability from the bivariate GREML, using SNPs common between two ancestries. WB, OE, SAS, AFR and MA indicates White British, Other European, South Asian, African, and Mixed ancestry cohorts.

**Supplementary Table 12: The estimated cross-ancestry genetic correlation (SE) for HDL-cholesterol**

|  | **Cohort1/**  **White British**  **(n=25,812)** | **Cohort2/**  **Other European**  **(n=22,368)** | **Cohort3/**  **South Asian**  **(n=5,063)** | **Cohort4/**  **African**  **(n=5,146)** |
| --- | --- | --- | --- | --- |
| **Cohort1/**  **White British** |  | **1.135 (0.302)**  **P= 6.55e-01**  $h_{c}^{2} (WB)$= 0.046 (0.012)  $h_{c}^{2} (OE)$= 0.050 (0.016) | **1.029 (0.237)**  **P= 9.02e-01**  $h_{c}^{2} (WB)$= 0.038 (0.011)  $h_{c}^{2} (As)$= 0.305 (0.056) | **0.488 (0.323)**  **P=1.13e-01**  $h_{c}^{2} (WB)$= 0.034 (0.010)  $h_{c}^{2} (Af)$= 0.191 (0.054) |
| **Cohort2/**  **Other European** |  |  | **1.003 (0.288)**  **P=9.92e-01**  $h_{c}^{2} (OE)$= 0.042 (0.015)  $h_{c}^{2} (As)$= 0.294 (0.056) | **0.652 (0.346)**  **P= 3.15e-01**  $h_{c}^{2} (OE)$= 0.045 (0.014)  $h_{c}^{2} (Af)$= 0.184 (0.054) |
| **Cohort3/**  **South Asian** |  |  |  | **0.375 (0.273)**  **P= 2.21e-02**  $h_{c}^{2} (As)$=0.257 (0.052)  $h_{c}^{2} (Af)$= 0.185 (0.055) |
| **Cohort4/**  **African** |  |  |  |  |

P is the *p*-value based on Wald’s test statistics for the null hypothesis of the estimated cross ancestry genetic correlation $r_{g}$=1 (i.e., a two-sided test). $h_{c}^{2}$ is the estimated SNP-based heritability from the bivariate GREML, using SNPs common between two ancestries. WB, OE, SAS, AFR and MA indicates White British, Other European, South Asian, African, and Mixed ancestry cohorts.

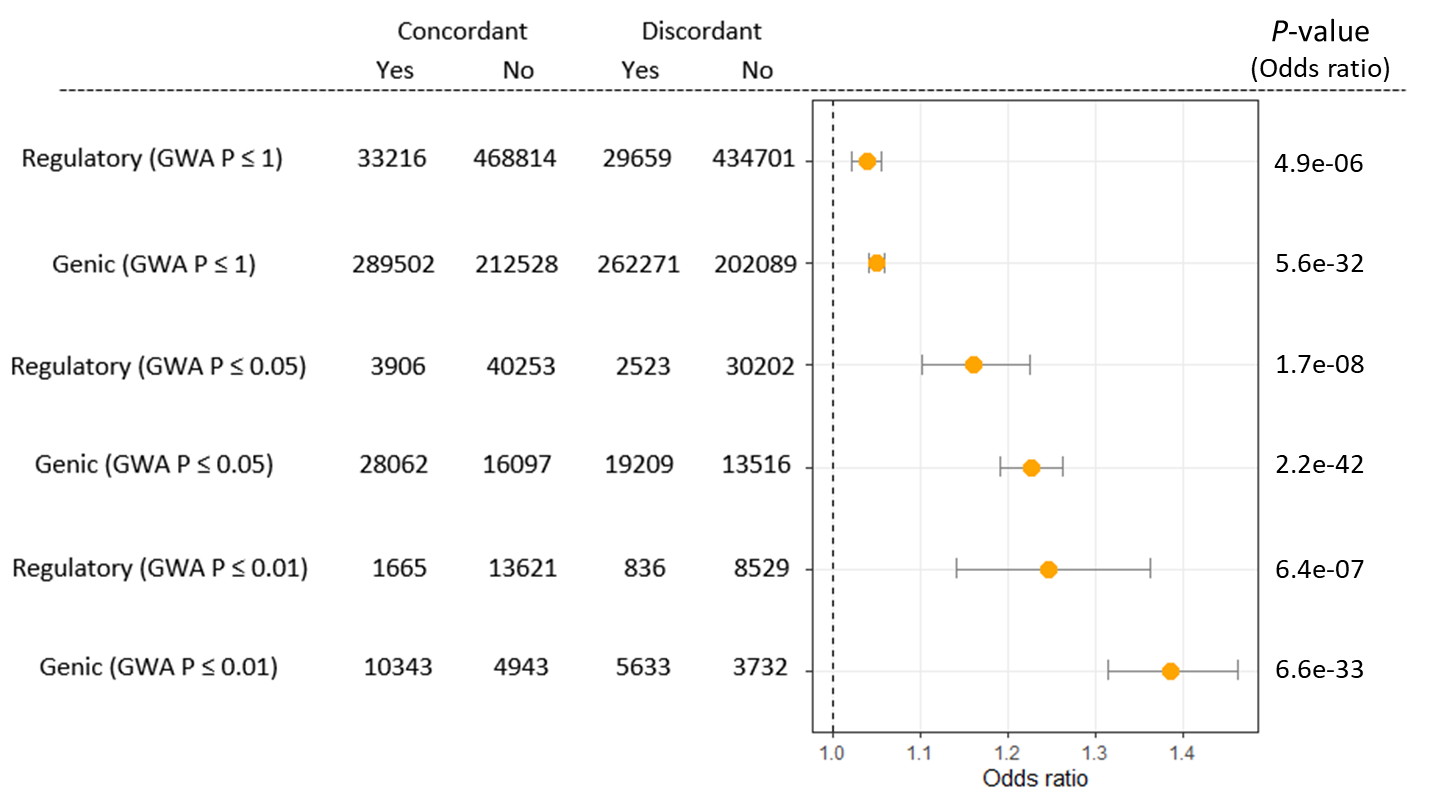

**Supplementary Figure 1: A forest plot with odds ratios indicating that concordant SNPs are more frequently found in the regulatory or genic region.** This analysis is for HDL-cholesterol phenotypes. Error bar represents 95% confidence intervals. The *p*-value of odds ratio indicates to be significantly different from 1. For regulatory or genic region, a genome-wide association (GWA) *p*-value threshold ≤1, 0.05 or 0.01 was used to select a set of concordant and discordant SNPs using UK Biobank GWAS summary statistics for HDL-cholesterol.

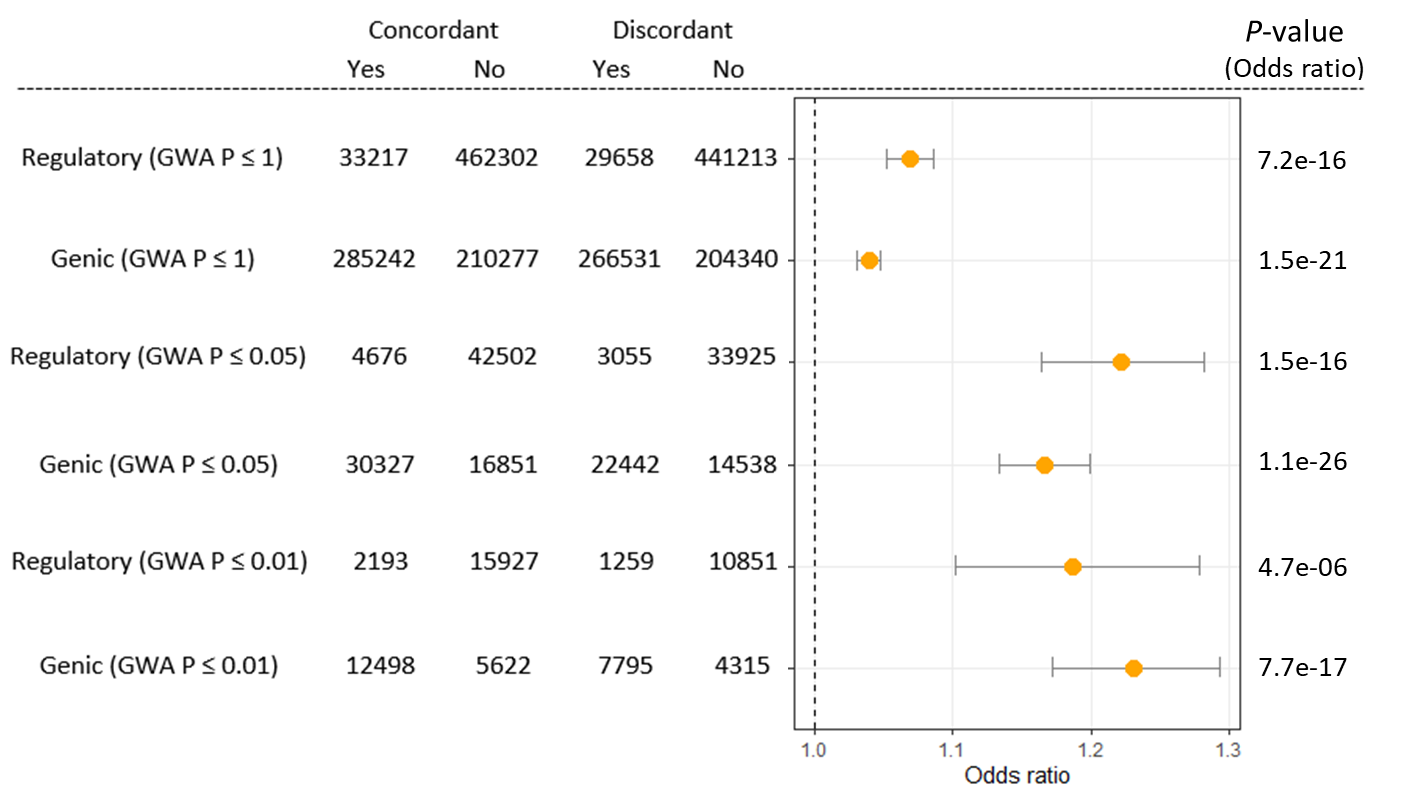

**Supplementary Figure 2: A forest plot with odds ratios indicating that concordant SNPs are more frequently found in the regulatory or genic region.** This analysis is for LDL-cholesterol phenotypes. Error bar represents 95% confidence intervals. The *p*-value of odds ratio indicates to be significantly different from 1. For regulatory or genic region, a genome-wide association (GWA) *p*-value threshold ≤1, 0.05 or 0.01 was used to select a set of concordant and discordant SNPs using UK Biobank GWAS summary statistics for LDL-cholesterol.

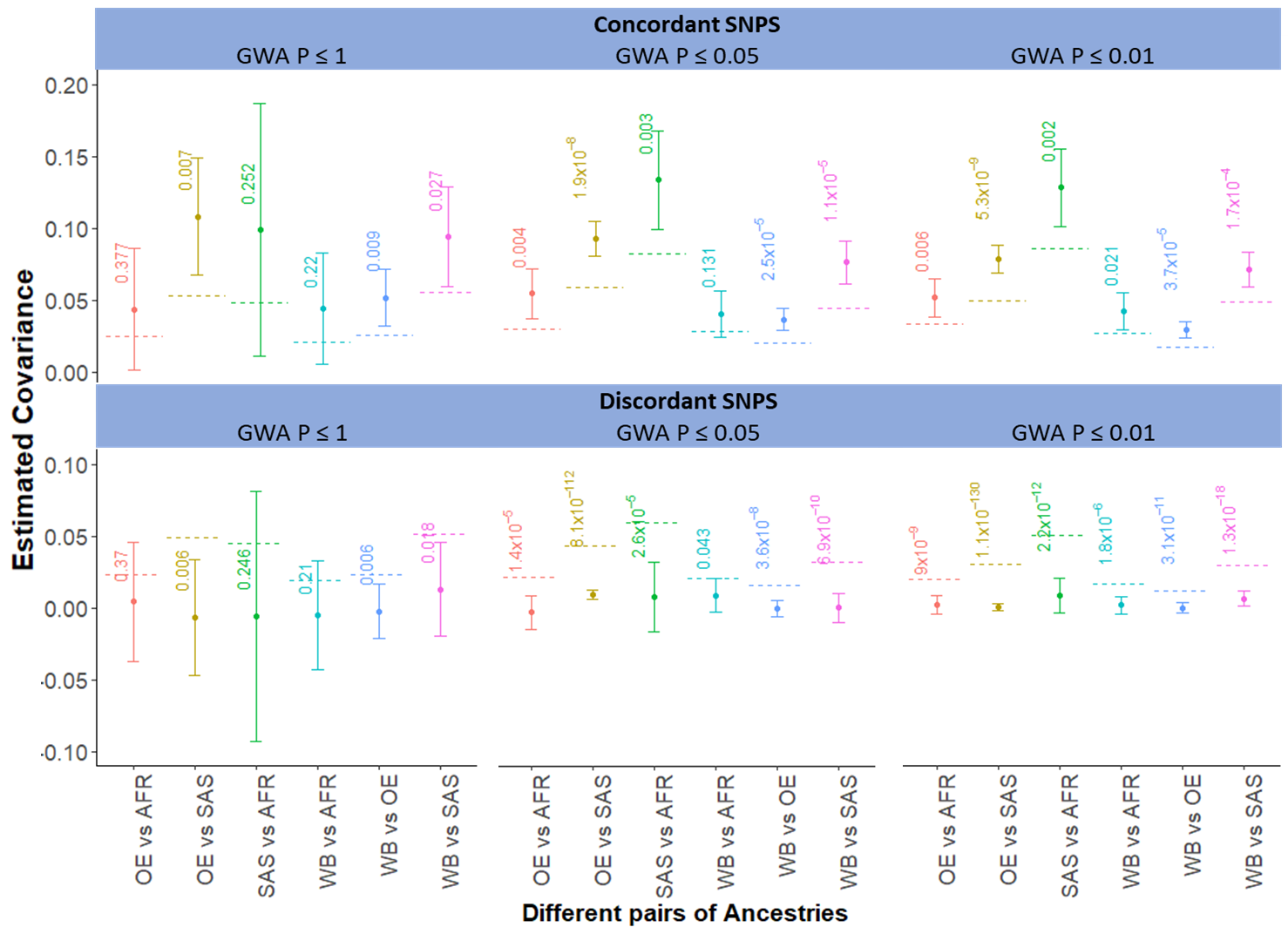

**Supplementary Figure 3: Estimated covariance for concordant and discordant SNPs for HDL-Cholesterol.** Concordant and discordant SNPs were derived from the comparison of SNP effects between two independent GWAS datasets of UK Biobank and BBJ. In this analysis, a set of SNPs with a genome-wide association (GWA) *p*-value < 1, 0.05 or 0.01 was used. The GWA p-values from UK Biobank GWAS for HDL-cholesterol were used. The main bars represent estimated cross-ancestry genetic covariance using the set of genome-wide SNPs, and the error bars indicate 95% confidence intervals. The dash line indicates the expected genetic covariance, assuming all SNPs contribute equally to the genetic covariance, i.e., the expected genetic covariance = the estimated total genetic covariance $\times$ the proportion of number of concordant SNPs, where the estimated total genetic covariance is based on all the SNPs including both concordant and discordant SNPs. The value with each bar indicates a *p*-value testing the null hypothesis that the estimated genetic covariance is not significantly different from the expectation. WB = White British, OE = Other European, SAS = South Asian, AFR = African.

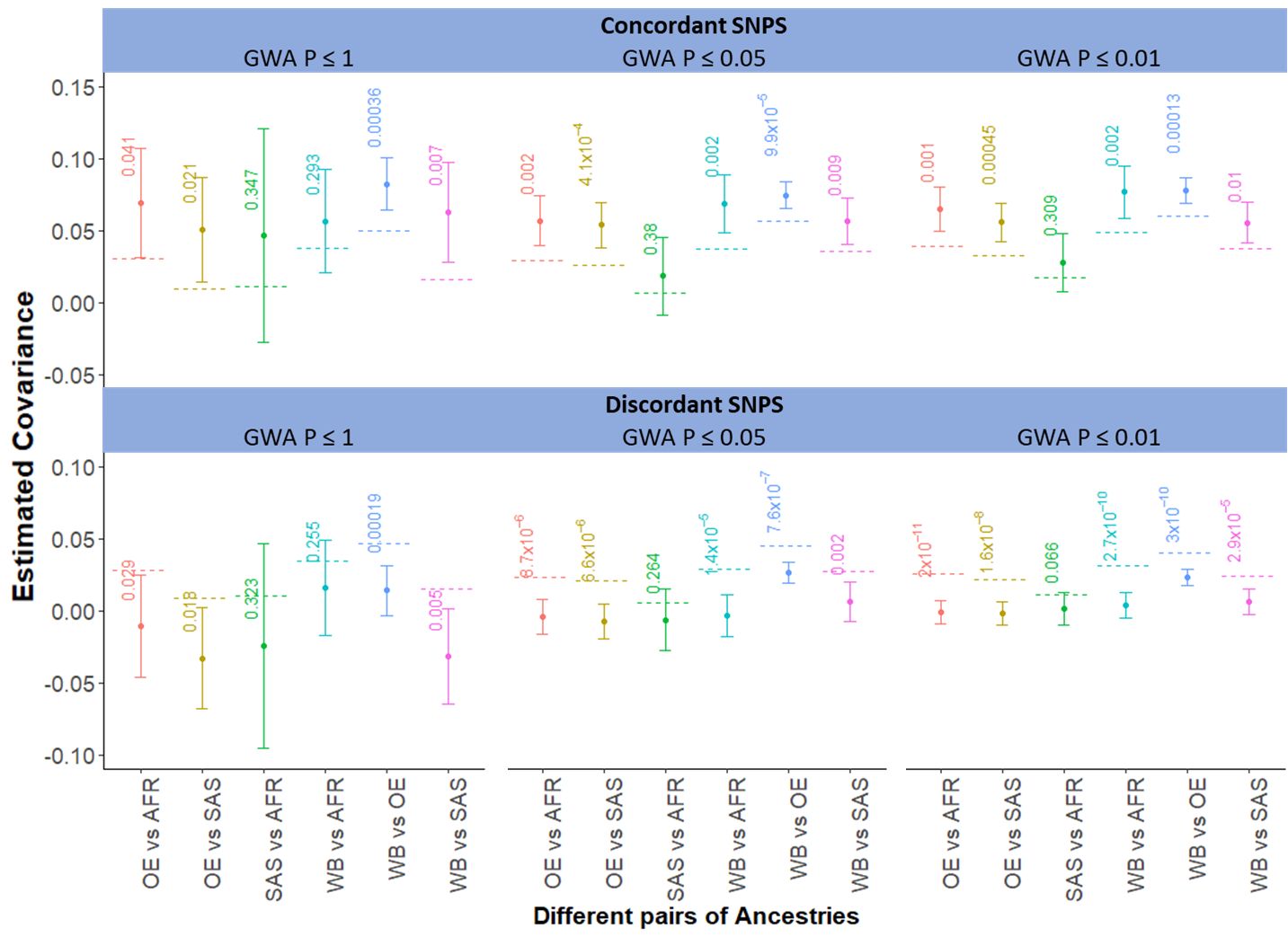

**Supplementary Figure 4:** **Estimated covariance for concordant and discordant SNPs for LDL-Cholesterol.** Concordant and discordant SNPs were derived from the comparison of SNP effects between two independent GWAS datasets of UK Biobank and BBJ. In this analysis, a set of SNPs with a genome-wide association (GWA) *p*-value < 1, 0.05 or 0.01 was used. The GWA p-values from UK Biobank GWAS for LDL-cholesterol were used. The main bars represent estimated cross-ancestry genetic covariance using the set of genome-wide SNPs, and the error bars indicate 95% confidence intervals. The dash line indicates the expected genetic covariance, assuming all SNPs contribute equally to the genetic covariance, i.e., the expected genetic covariance = the estimated total genetic covariance $\times$ the proportion of number of concordant SNPs, where the estimated total genetic covariance is based on all the SNPs including both concordant and discordant SNPs. The value with each bar indicates a *p*-value testing the null hypothesis that the estimated genetic covariance is not significantly different from the expectation. WB = White British, OE = Other European, SAS = South Asian, AFR = African.

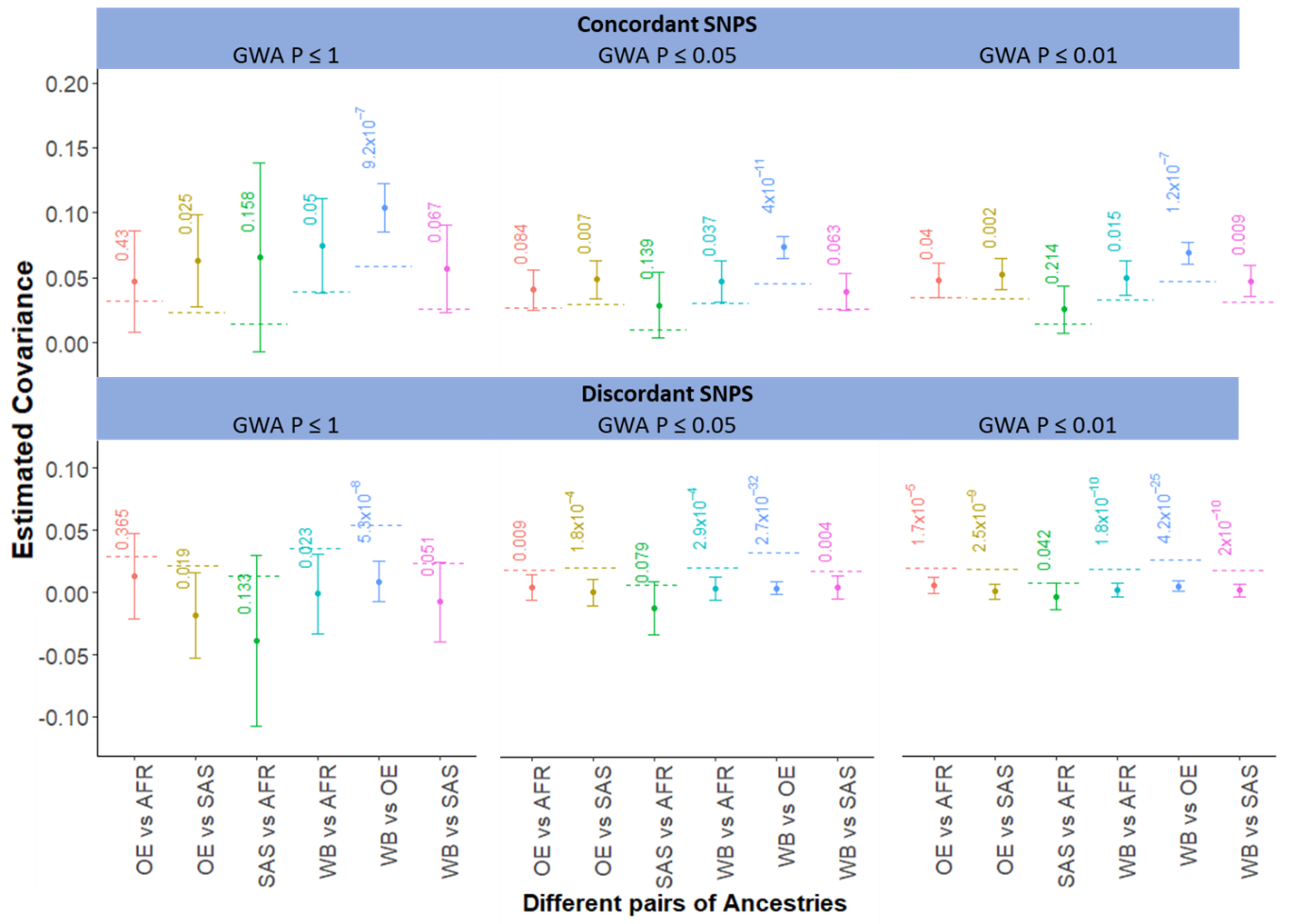

**Supplementary Figure 5: Estimated covariance for concordant and discordant SNPs for total-cholesterol.** Concordant and discordant SNPs were derived from the comparison of SNP effects between two independent GWAS datasets of UK Biobank and BBJ. In this analysis, a set of SNPs with a genome-wide association (GWA) *p*-value < 1, 0.05 or 0.01 was used. The GWA p-values from Biobank Japan GWAS for total-cholesterol were used. The main bars represent estimated cross-ancestry genetic covariance using the set of genome-wide SNPs, and the error bars indicate 95% confidence intervals. The dash line indicates the expected genetic covariance, assuming all SNPs contribute equally to the genetic covariance, i.e., the expected genetic covariance = the estimated total genetic covariance $\times$ the proportion of number of concordant SNPs, where the estimated total genetic covariance is based on all the SNPs including both concordant and discordant SNPs. The value with each bar indicates a *p*-value testing the null hypothesis that the estimated genetic covariance is not significantly different from the expectation. WB = White British, OE = Other European, SAS = South Asian, AFR = African.

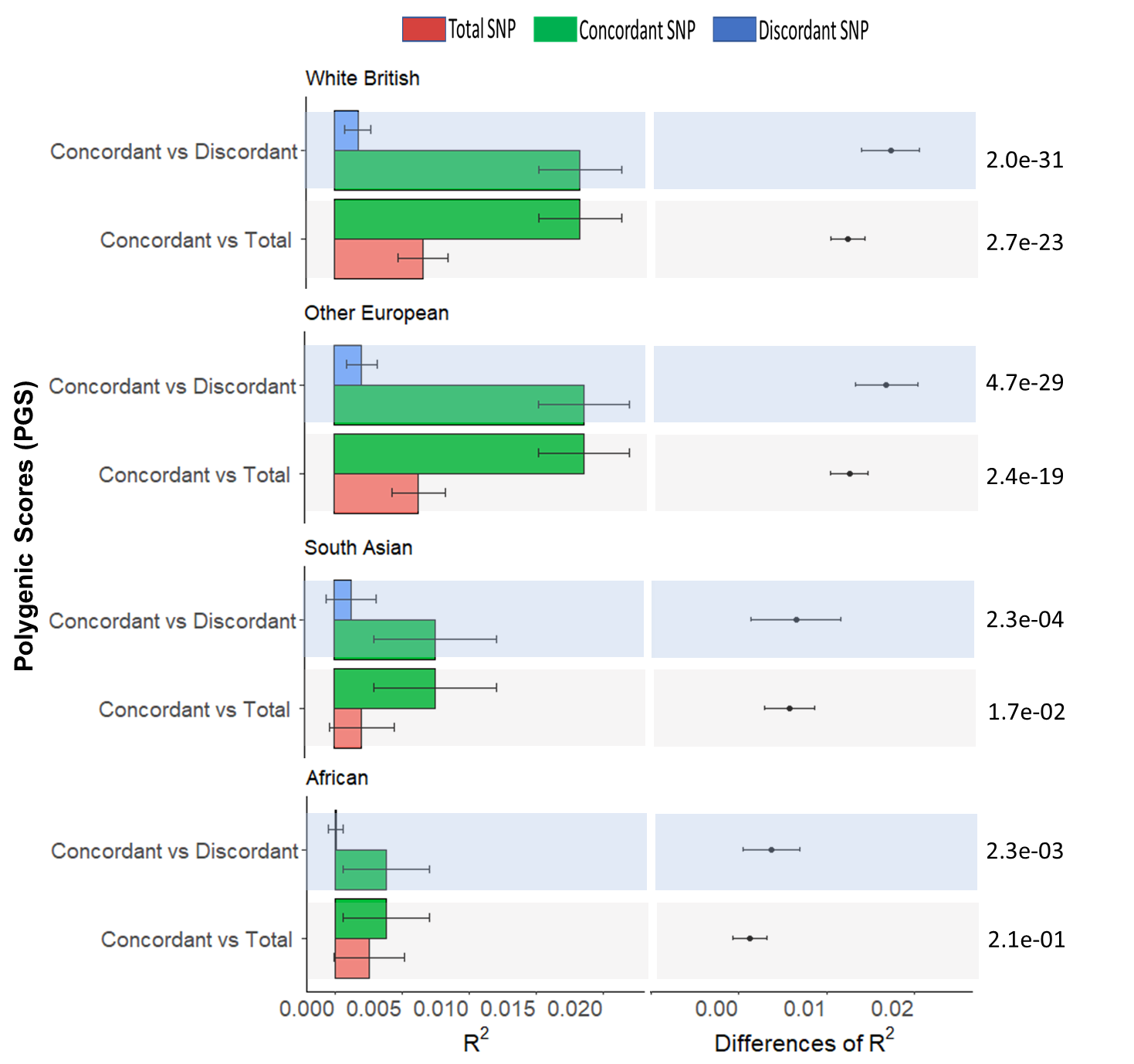
**Supplementary Figure 4: The predictive ability (**$\boldsymbol{R}^{\boldsymbol{2}}$**) of polygenic risk scores for total-cholesterol when using the set of concordant, discordant, or total SNPs for cross-ancestry risk predictions.**

Biobank Japan GWAS was used as discovery dataset (n=128,305, while target datasets were other European (n=26,457), south Asian (n=6,199) and African (n=6,179).

**Left panels:** The main bars represent $R^{2}$ values and error bars correspond to 95% confidence interval.

**Right panels:** Dot points represents the differences between $R^{2}$ values, and error bars correspond to 95% confidence intervals of the differences. The *p*-value in each bar indicates that the differences of **(**$\boldsymbol{R}^{\boldsymbol{2}}$**)** are significantly different from zero.

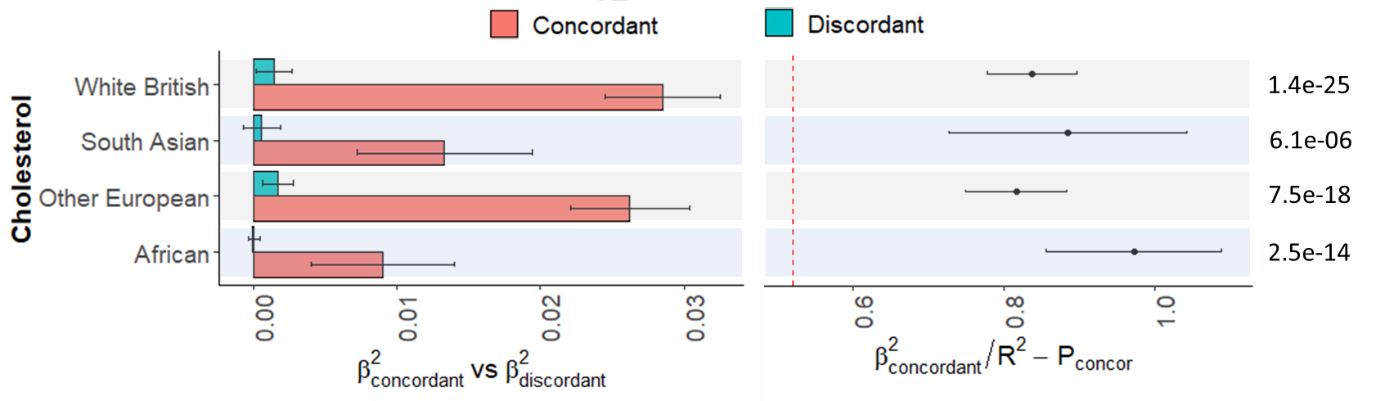

**Supplementary Figure 5: PGS-based genomic partitioning method to assess if the predictive ability is enriched for the concordant SNPs for Total-cholesterol.** Here, $P_{concor}=$0.52 is the expectation for the proportion of concordant SNPs based on this annotation.

**Left panel:** The main bars represent squared regression coefficients attributable to SNPs for the concordant ($\hat{\beta}_{concordant}^{2}$) and discordant SNPs list ($\hat{\beta}_{discordant}^{2}$), and error bars correspond 95% confidence intervals when predicting white British, other European, South Asian, and African.

**Right panel:** Dot points represent the difference between the observed and expected proportions ($\frac{\hat{\beta}_{concordant}^{2}}{R^{2}}-P_{concor}$) and error bars indicate 95% confidence intervals of the difference. The *p*-value in each bar indicates that the difference between the observed and expected proportions ($\frac{\hat{\beta}_{concordant}^{2}}{R^{2}}-P_{concor}$) are significantly different from zero.

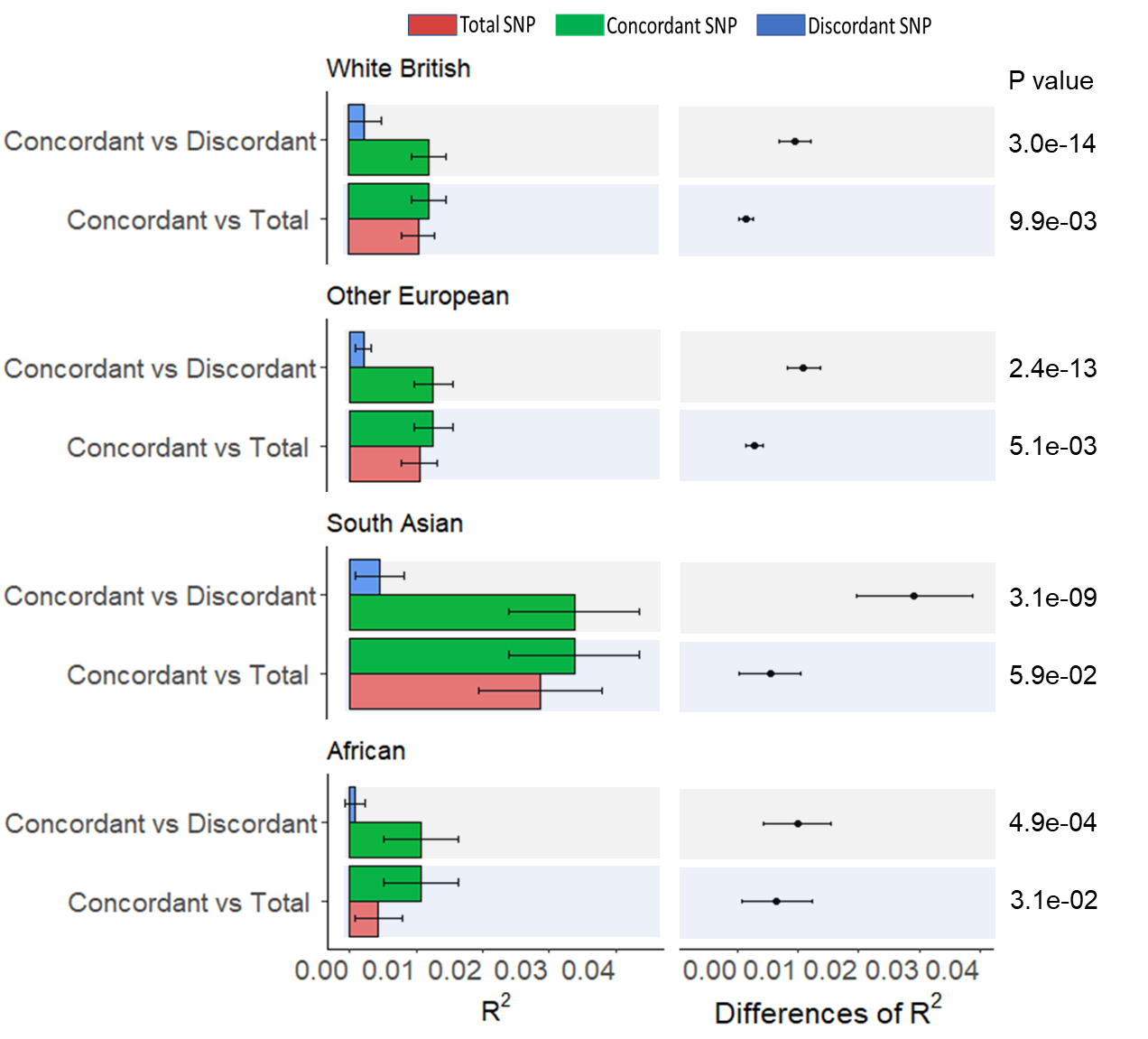

**Supplementary Figure 6:** **The predictive ability (**$\boldsymbol{R}^{\boldsymbol{2}}$**) of polygenic risk scores for HDL-cholesterol when using the set of concordant, discordant, or total SNPs for cross-ancestry risk predictions.** UK Biobank GWAS was used as the discovery dataset (n= 258,792), while target datasets were other European (n=26,457), south Asian (n=6,199) and African ancestry (n=6,179).

**Left panels:** The main bars represent $R^{2}$ values and error bars correspond to 95% confidence interval.

**Right panels:** Dot points represent the differences between $R^{2}$ values, error bars correspond to 95% confidence intervals of the differences, and *p*-values indicate that the differences of $R^{2}$ are significantly different from zero (null hypothesis). P-values was estimated using an R-package named r2redux^3^ based on Wald’s test statistics.

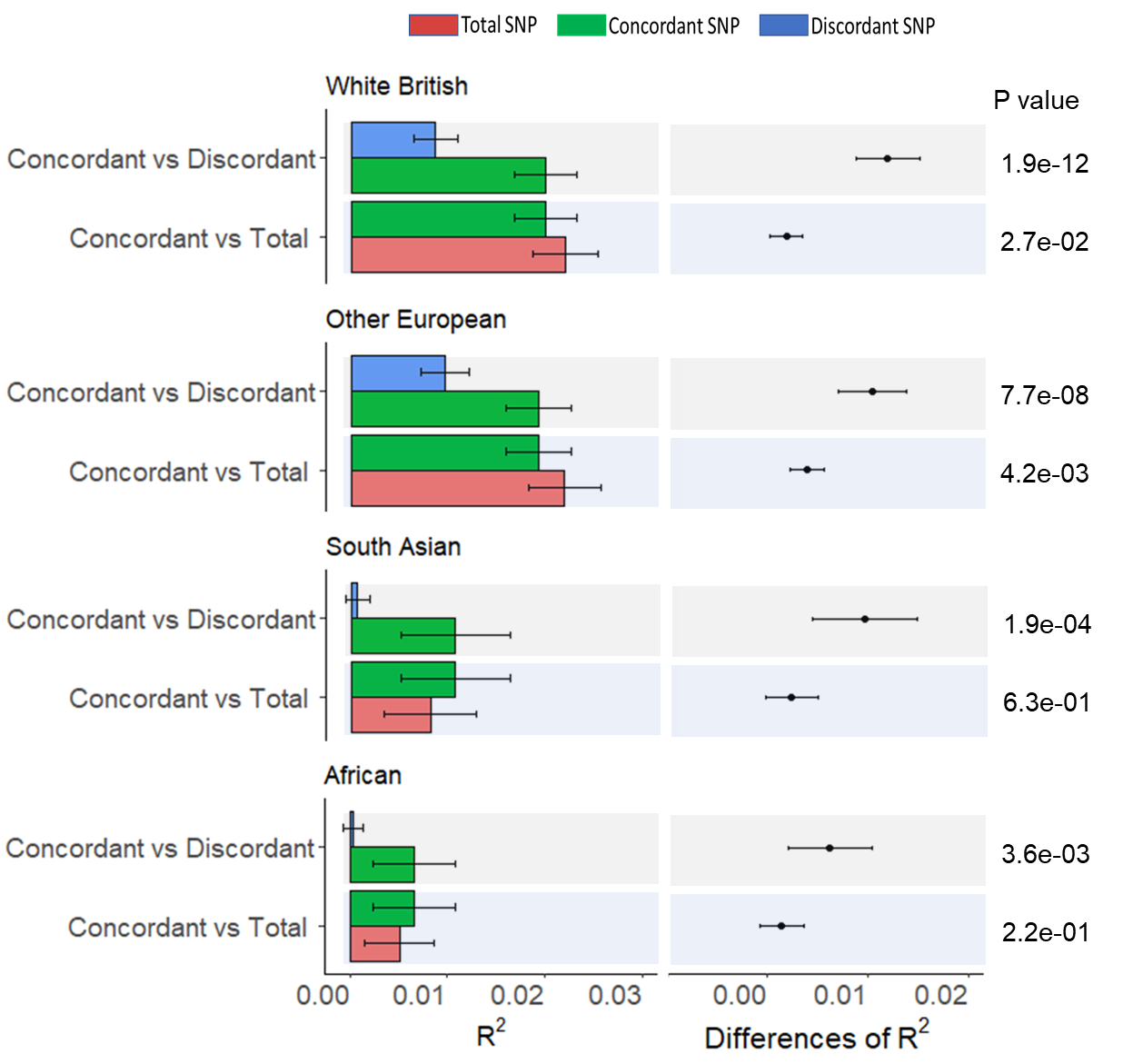

**Supplementary Figure 7: The predictive ability (**$\boldsymbol{R}^{\boldsymbol{2}}$**) of polygenic risk scores for LDL-cholesterol when using the set of concordant, discordant, or total SNPs for cross-ancestry risk predictions.** UK Biobank GWAS was used as the discovery dataset (n= 258,792), while target datasets were other European (n=26,457), south Asian (n=6,199) and African ancestry (n=6,179).

**Left panels:** The main bars represent $R^{2}$ values and error bars correspond to 95% confidence interval.

**Right panels:** Dot points represent the differences between $R^{2}$ values, error bars correspond to 95% confidence intervals of the differences, and *p*-values indicate that the differences of $R^{2}$ are significantly different from zero (null hypothesis). P-values was estimated using an R-package named r2redux^3^ based on Wald’s test statistics.

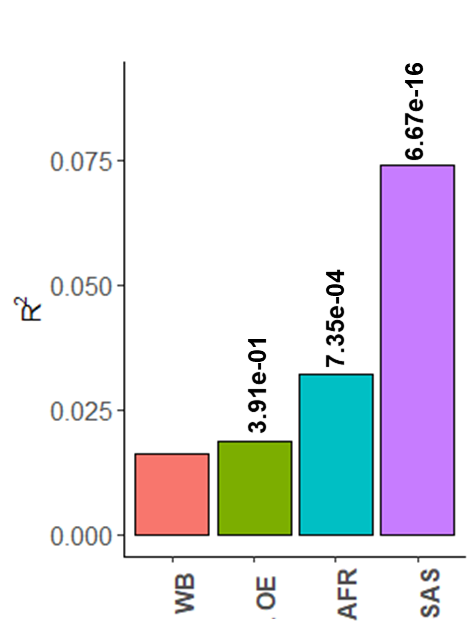

**Supplementary Figure 8*.* The predictive ability (**$\mathbf{R}^{\mathbf{2}}$**) of PGS for HDL-cholesterol within-ancestry prediction (WB) and cross-ancestry predictions (OE, AFR, and SAS).** Cross-ancestry $R^{2}$is estimated using total SNPs (both concordant and discordant SNPs) and clump-and-threshold (C + T) based PRS method (PRSice)^4^. WB, OE, SAS, and AFR indicates White British, Other European, South Asian, and African ancestry cohorts, which were used as target datasets (n = 30,000, 26,457, 6,199 and 6,179). PGS were constructed based on SNP effects estimated using 258,000 white British individuals as the discovery dataset that are independent from those target datasets. The value above each bar is the p-value indicating if the R^2^ of cross-ancestry prediction (OE, AFR or SAS) is significantly different from that of within-ancestry prediction (WB). P-values between two independent PGS was estimated following Momin et al.^3^.
